# A kidney-conditioned urinary peptidomic biological ageing clock and its association with all-cause mortality and age-related health outcomes: a retrospective multicohort development and validation study

**DOI:** 10.64898/2026.09.01.26361640

**Authors:** Sajjad Biglari, Mayra Alejandra Jaimes-Campos, Justyna Siwy, Agnieszka Latosinska, Harald Mischak, Tim S Nawrot, Jan A Staessen, Dries S Martens, Miroslaw Banasik

## Abstract

**Background:** Urine is non-invasive and molecularly rich, but filtered and shaped by kidney function. We aimed to develop a kidney-conditioned urinary peptidomic ageing clock and relate its age acceleration to mortality and incident disease.

**Methods:** In this retrospective multicohort study, urinary peptides were measured by capillary electrophoresis–mass spectrometry. A urinary peptidomic ageing clock (UPBioAge) was developed in a kidney function-preserved derivation cohort (n=1,811), then conditioned on estimated glomerular filtration rate (eGFR) and albuminuria by Filtrate-Aware Calibration in a separate kidney-diverse cohort (n=7,797), giving k-UPBioAge. Kidney-conditioned age acceleration (k-UPBioAgeAcc) was related to all-cause mortality and incident disease in a clinically enriched follow-up cohort (n=7,469; 2,854 [38·2%] women; urine collected 1999– 2019; 625 deaths; median follow-up 3·95 years [IQR 2 39–5 58]) using Cox models adjusted for age, sex, comorbidities, body-mass index, blood pressure, and eGFR.

**Findings:** k-UPBioAge had a calibrated holdout mean absolute error of 4·94 years, and 5·44 years in two validation cohorts. Per SD, k-UPBioAgeAcc was associated with all-cause mortality (hazard ratio 1·47, 95% CI 1 34–1 62), incident coronary artery disease (1 44, 1 27–1 64), heart failure (1 29, 1 15–1 44), and chronic kidney disease progression (1 39, 1 08–1 78).

**Interpretation:** Urinary ageing clocks may need conditioning on kidney function. So conditioned, a single urine sample carried prognostic information beyond age and clinical risk factors; after prospective validation, it could support risk stratification and serve as a response measure in trials targeting ageing.

**Funding:** EU Horizon Europe Marie Skłodowska-Curie Actions (PICKED); German Federal Ministry of Education and Research (ERA PerMed SIGNAL); COST Actions PerMediK and KidneyHealth.

**Research in context:** *Evidence before this study:* We searched PubMed for studies published in English up to August 6, 2026, using terms covering the urinary peptidome combined with biological age and with kidney function; all 35 records identified were screened in full. Most published ageing clocks are based on DNA methylation or serum or plasma proteomics, and we identified a single urinary peptidomic ageing clock, the capillary electrophoresis–mass spectrometry (CE–MS)-based UPP-age, which has been associated with chronological age, disease, and mortality. To our knowledge, no urinary ageing clock has treated urine as a filtered biofluid whose age signal is shaped by kidney function, or been conditioned on it.

*Added value of this study:* A urinary peptidomic ageing clock, conditioned on kidney function, was associated with all-cause mortality and with incident coronary artery disease, heart failure, and chronic kidney disease progression beyond established clinical risk factors, and was higher in people with multimorbidity. Without conditioning, the same clock rated people with the best-preserved kidney function as the most biologically aged, and its association with mortality weakened as kidney function declined until it was lost in advanced chronic kidney disease. No outcome data were used in developing the clock or the calibration.

*Implications of all the available evidence:* A single urine sample, collected without a needle, can provide a measure of biological ageing that is associated with death and incident disease, provided it is read in the context of kidney function. After prospective validation, such a clock could help identify older and multimorbid patients whose risk exceeds what their age and clinical profile suggest, and serve as a non-invasive response measure in trials of interventions targeting ageing. Any urinary ageing marker may need conditioning on kidney function, and the same principle, treating the producing organ’s function as part of the assay, may apply to other organ-derived biofluids such as saliva and cerebrospinal fluid.

## Introduction

Ageing is the progressive loss of cellular and physiological function, leading to increased susceptibility to disease and death.^1^ Multiple underlying biological processes, referred to as the hallmarks of ageing, shape individual ageing trajectories.^2^ Molecular ageing clocks capture differences in biological age between people of the same chronological age and could support risk stratification and the monitoring of interventions targeting healthy ageing.^3^

Current ageing clocks based on DNA methylation,^3^ transcriptomics,^4^ proteomics,^5^ and metabolomics^6^ have mostly been developed from blood. Urine represents an attractive but underexplored matrix for studying biological ageing, as the urinary peptidome is biologically rich, with more than 20,000 reproducibly detectable endogenous peptide features,^7^ and reflects both renal and extra-renal pathophysiological processes.^8^ Urine can be collected non-invasively, repeatably, and at low cost, and the urinary peptidome is stable under standardised collection, processing, and storage.^9^

Previous studies have identified age-associated peptides in urine.^10^ A urinary peptidomic ageing clock (UPP-age) was associated with mortality in the general population.^11^ However, urine is a filtered biofluid, and age-related differences in glomerular filtration and tubular handling, together with albuminuria-associated peptide changes, can alter the measured peptidome.^12,13^ Studies in chronic kidney disease (CKD) show that urinary collagen fragments and peptidomic classifiers track kidney function decline and histological fibrosis.^14^ How far this kidney-related component confounds biological age prediction has not been addressed.

We therefore developed a urinary peptidomic ageing clock in people with preserved kidney function, conditioned it on estimated glomerular filtration rate (eGFR) and urinary albumin-to-creatinine ratio (UACR) in an independent cohort spanning the full range of kidney function, and tested whether the resulting age acceleration predicts all-cause mortality and incident cardiovascular and kidney disease in a clinically enriched cohort, beyond established clinical risk factors.

## Methods

### Study design and participants

The cohorts of this retrospective multicohort study are summarised in Table 1 and the analytical flow in Figure 1A–C, with sample counts and the detailed workflow in Supplementary Figure S1. Full eligibility criteria and cohort assembly are given in Supplementary Methods S1. The derivation cohort comprised 1,811 participants from the Human Urinary Proteome Database,^7^ aged 18 years or older with normoalbuminuria (UACR <30 mg/g; Kidney Disease: Improving Global Outcomes [KDIGO] category A1) and preserved kidney function, with eGFR (Chronic Kidney Disease Epidemiology Collaboration [CKD-EPI] equation) interpreted in an age-adapted way because filtration declines physiologically with age, as proposed in age-adapted definitions of CKD.^15,16^ Participants with kidney disease, cancer, COVID-19, heart failure (HF), coronary artery disease (CAD) and severe hypertension (≥180/120 mm Hg) were excluded, so that the clock would be less likely to learn disease-related peptide patterns. Participants were recorded as healthy controls in the source database. The cohort was split 80/20 into training (n=1,449) and internal holdout (n=362) sets, stratified by age and sex. Reporting follows the STROBE and TRIPOD+AI guidelines.

**Figure 1.**
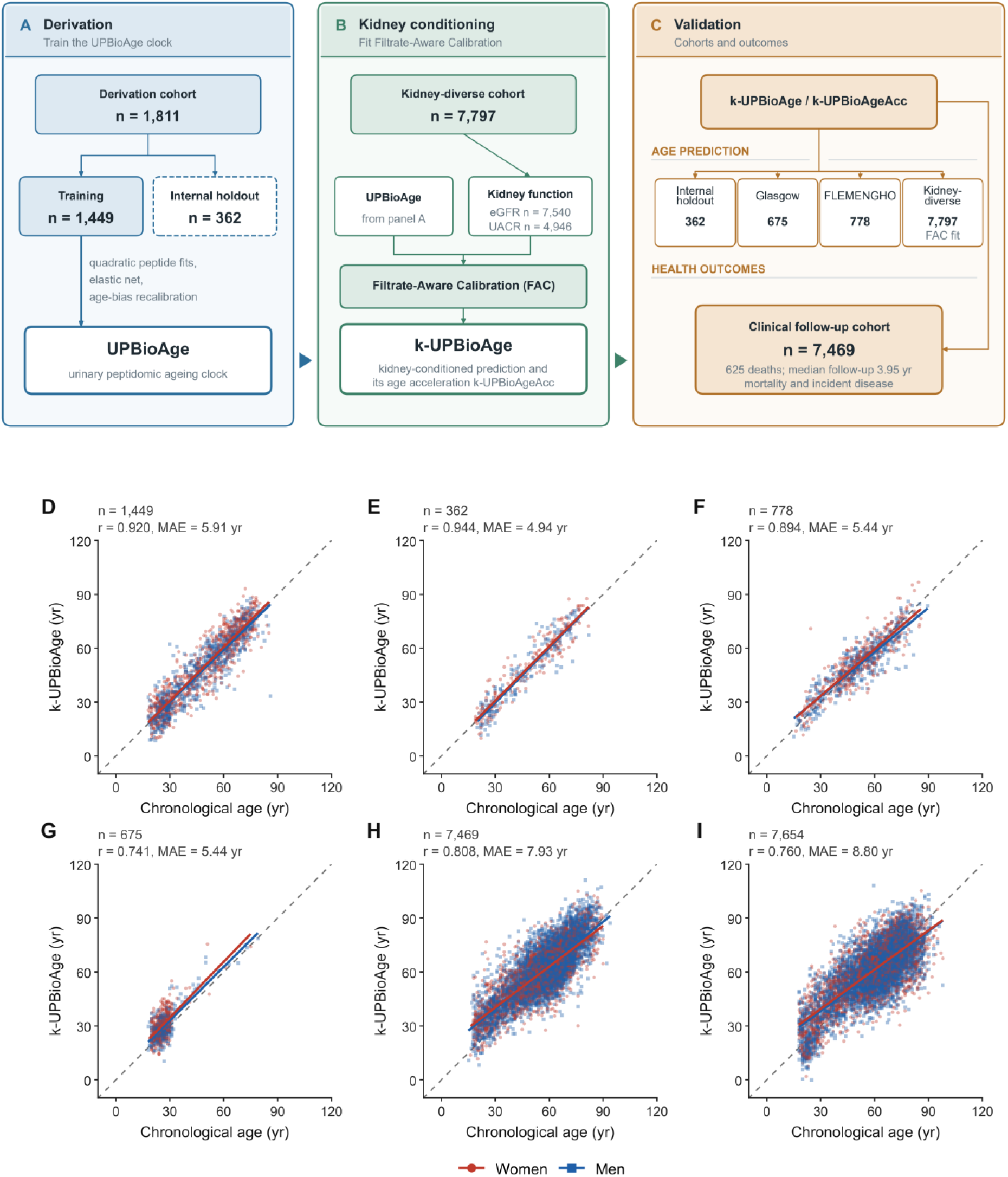
Study design and age prediction. A–C, Study design. A, The clock is trained on 1,449 of 1,811 derivation participants, with 362 held out. B, Filtrate-Aware Calibration (FAC) is fitted in the kidney-diverse cohort (n=7,797; measured eGFR in 7,540 and UACR in 4,946), yielding k-UPBioAge and its age acceleration k-UPBioAgeAcc. C, Validation cohorts and outcomes. D–I, k-UPBioAge against chronological age in D, training (n=1,449); E, internal holdout (n=362); F, FLEMENGHO (n=778); G, Glasgow (n=675); H, clinical follow-up (n=7,469); and I, the kidney-diverse cohort (n=7,654 with age and sex recorded). Red circles, women; blue squares, men; solid lines are sex-specific least-squares fits and the dashed line is identity. Pearson r and MAE are for the cohort as a whole. The kidney-diverse cohort in I is the set in which FAC was fitted; it shares no participants with any other panel, but it is not independent of the kidney-conditioning step. No validation participant contributed to clock derivation or to FAC modelling; 775 of the 778 FLEMENGHO participants also belong to the clinical follow-up cohort. eGFR, estimated glomerular filtration rate; FAC, Filtrate-Aware Calibration; FLEMENGHO, Flemish Study on Environment, Genes and Health Outcomes; MAE, mean absolute error; UACR, urinary albumin-to-creatinine ratio; k-UPBioAge, kidney-conditioned urinary peptidomic biological age; k-UPBioAgeAcc, kidney-conditioned urinary peptidomic biological age acceleration.

**Table 1.** Baseline characteristics of the analytic cohorts.

|  | n | Age, yr | Women, n (%) | eGFR, mL/min/1.73 m<sup>2</sup> | UACR, mg/g | BMI, kg/m<sup>2</sup> | SBP, mmHg | DBP, mmHg |
| --- | --- | --- | --- | --- | --- | --- | --- | --- |
| **Biological age-clock** |  |  |  |  |  |  |  |  |
| *Derivation cohort* |  |  |  |  |  |  |  |  |
| Training | 1,449 | 48.5 (17.9) | 784 (54.1%) | 91.6 (32.4) | 8.6 (6.5) | 28.7 (6.1) | 131.2 (17.4) | 77.9 (8.7) |
| Internal holdout | 362 | 48.4 (18.0) | 196 (54.1%) | 89.8 (25.6) | 9.3 (6.5) | 29.5 (6.2) | 132.6 (15.6) | 79.2 (9.1) |
| *Validation cohorts* |  |  |  |  |  |  |  |  |
| Clinical follow-up | 7,469 | 57.0 (15.3) | 2,854 (38.2%) | 78.8 (28.7) | 67.0 (380.6) | 28.5 (5.4) | 133.3 (17.9) | 78.1 (10.1) |
| Glasgow | 675 | 26.3 (6.4) | 368 (54.5%) | 123.0 (24.9) | 30.8 (312.5) | 25.4 (5.9) | 119.3 (13.1) | 78.6 (9.3) |
| FLEMENGHO | 778 | 51.0 (15.8) | 395 (50.8%) | 82.7 (17.4) | 15.1 (32.0) | 26.3 (4.0) | 129.0 (17.5) | 79.5 (9.7) |
| **Surrogate + FAC** |  |  |  |  |  |  |  |  |
| Kidney-diverse cohort | 7,797 | 59.6 (16.4) | 2,578 (33.6%) | 78.8 (46.6) | 772.3 (1,741.2) | 28.3 (5.8) | 131.3 (20.9) | 73.4 (13.4) |

Data are mean (SD) or n (%). Sex was not recorded for 117 participants of the kidney-diverse cohort; its percentage of women is based on the 7,680 with sex recorded. UACR is strongly right-skewed, so its mean exceeds its median in every cohort and its SD is correspondingly large. Available data for eGFR and UACR were as follows: 77.6% and 61.3% for the training cohort, 79.6% and 64.4% for the internal holdout, 100% and 33.2% for clinical follow-up cohort, 99.9% and 99.7% for Glasgow cohort, 100% and 92.0% for the FLEMENGHO cohort, and 96.7% and 63.4% for kidney-diverse cohort. Of the 778 FLEMENGHO participants, 775 also belong to the clinical follow-up cohort. Abbreviations: FAC: Filtrate-Aware Calibration; FLEMENGHO: Flemish Study on Environment, Genes and Health Outcomes; eGFR: estimated glomerular filtration rate; UACR: urinary albumin-to-creatinine ratio; BMI: body-mass index; SBP: systolic blood pressure; DBP: diastolic blood pressure; SD: standard deviation; yr: years.

Validation used 675 participants (604 from South African studies) analysed in an independent laboratory in Glasgow, UK; all other samples were analysed in the reference laboratory in Hanover, Germany. The Flemish Study on Environment, Genes and Health Outcomes (FLEMENGHO) provided further validation, with urine collected in 2005–2011 from 778 participants, 775 of whom are also in the clinical follow-up cohort.^11^ Adverse health outcomes were evaluated in a clinically enriched cohort of 7,469 participants with 625 deaths and a median follow-up of 3 95 years (IQR 2 39–5 58), with all-cause mortality as the primary endpoint; its mortality data were analysed previously with outcome-trained classifiers.^17^ The cohort combined 17 contributing studies with prospective follow-up, predominantly in diabetes, kidney disease, cardiovascular disease, and hypertension. Secondary endpoints were incident CAD, HF, CKD progression (eGFR decline ≥40%), and a composite cardiovascular endpoint, the first of either a CAD or an HF event. Endpoints were ascertained within the contributing studies under their own adjudicated definitions, independently of the peptidomic measurements; no outcome data were used by the investigators during clock development or peptide selection. Contributing studies, follow-up time, and incidence per endpoint are in Supplementary Tables S1 and S2.

### Procedures

Urine (midstream second-void) samples were processed by capillary electrophoresis–mass spectrometry (CE–MS) as previously described,^18^ with peptide sequences assigned by CE– MS/MS and liquid chromatography–tandem mass spectrometry (LC–MS/MS) (Supplementary Methods S2). The complete panel comprised 12,019 sequenced urinary peptides, each mapped to a parent protein; intensities were natural log(x+1)-transformed, centred, and scaled. The urinary peptidomic biological age clock (UPBioAge) is a two-stage model followed by linear recalibration and a spline correction for regression to the mean across the 20–80 year range. The spline is a function of chronological age, evaluated at each participant’s own age whenever the clock is applied, so reported accuracy is calibrated rather than age-blind. Details are in Supplementary Methods S3.

UPBioAge was then conditioned on kidney function by Filtrate-Aware Calibration (FAC; Supplementary Methods S4). In a separate kidney-diverse cohort, the error of the linearly recalibrated prediction (before the age-dependent spline correction) was modelled as a smooth function of eGFR and UACR, and this correction was added to UPBioAge to give the kidney-conditioned prediction, k-UPBioAge. The correction applies only where eGFR is below 60 mL/min/1 73 m or UACR above 30 mg/g, with peptide-based estimates used for missing measurements; elsewhere the original prediction is kept. Kidney-conditioned age acceleration (k-UPBioAgeAcc) is the residual of k-UPBioAge on chronological age; a positive value means the urinary peptidome looks older than expected.

### Statistical analysis

Age-prediction performance was reported as mean absolute error (MAE), root mean squared error (RMSE), Pearson correlation, and coefficient of determination (R ). All-cause mortality was the primary endpoint, and its association with k-UPBioAgeAcc was evaluated by multivariable-adjusted Cox proportional hazards regression, with hazard ratios (HRs) per standard deviation (SD) increment and 95% Wald confidence intervals (CIs). A basic model included age and sex; a fully adjusted model added body-mass index (BMI), eGFR, mean arterial pressure, diabetes, cardiovascular disease, and hypertension. Proportional hazards for k-UPBioAgeAcc were assessed using scaled Schoenfeld residuals against log time. Where two HRs were compared directly, the difference in log-HRs was assessed by paired bootstrap. Quintile analyses are described in Supplementary Methods S5.

Prevalent multimorbidity was related to k-UPBioAgeAcc by linear regression, classifying participants by all observed combinations of diabetes, cardiovascular disease, kidney disease, and hypertension, with those free of all four as reference. Models were adjusted for chronological age, sex, BMI, and mean arterial pressure, and coefficients are expressed as the difference in k-UPBioAgeAcc in years with 95% confidence intervals. Sensitivity analyses of unconditioned age acceleration, adjustment for kidney-function measures, pathway over-representation, and model discrimination are described in Supplementary Methods S5–S8. Fairness was assessed as consistency of prediction accuracy and of the mortality association across sex, age group, KDIGO stage, albuminuria category, and comorbidity strata, with formal interaction tests. Sex was taken as recorded in the contributing studies (female or male); gender was not recorded. Ethnicity was not recorded in the source database, so performance across ancestry groups could not be examined.

### Ethics

This is a secondary analysis of fully anonymised data from previously published studies held in the Human Urinary Proteome Database. The Ethics Committee of Hannover Medical School, Germany, has waived the requirement for ethical approval for analyses of such fully anonymised data (No. 3116-2016). Ethical approval and written informed consent were obtained in each original recruitment study. No new samples were collected, consistent with the Declaration of Helsinki and the General Data Protection Regulation.

### Role of the funding source

The funders had no role in study design, data collection, data analysis, data interpretation, or writing of the report. All authors had full access to all the data in the study and had final responsibility for the decision to submit for publication.

## Results

Table 1 summarises the cohorts. The clinical follow-up cohort was older than the derivation cohort (mean age 57 0 [SD 15 3] against 48 5 [17 9] years) and clinically enriched: 54 8% of participants had diabetes, 41 3% hypertension, 29 6% kidney disease, and 18 2% cardiovascular disease (Supplementary Table S3). Of the 12,019 sequenced urinary peptides, 4,340 were detected in at least 40% of the 1,449 training participants, and 3,296 of these correlated with chronological age after Benjamini–Hochberg (BH) correction (Supplementary Figure S2). The final UPBioAge clock comprised 2,631 peptides from 685 parent proteins; the 1,683 (64 0%) collagen-derived peptides contributed 61 4% of the age estimate by model weight, dominated by the collagen type I α1 chain (COL1A1; 410 peptides, 14·6%) and the type III α1 chain (COL3A1; 283 peptides, 11·3%). Within each chain, fragments moved in both directions with age. The largest single contributor was a fragment of platelet endothelial cell adhesion molecule-1 (PECAM-1; CD31), and the leading non-collagen proteins were fibrinogen α (23 peptides) and the polymeric immunoglobulin receptor (22 peptides; Supplementary Table S4). All 2,631 peptides are listed in Supplementary Data 1 with their share of model weight. Pathway over-representation analysis identified 37 significantly enriched pathways involving collagen homeostasis, extracellular-matrix organisation, and integrin signalling; because the urinary peptidome consists of proteolytic fragments, enrichment of collagen synthesis terms indexes turnover rather than net synthesis (Supplementary Figure S3; Supplementary Table S5).

In the internal holdout cohort, k-UPBioAge explained 87 4% of the variation in chronological age, with a MAE of 4 94 years after the standard age-bias correction (Figure 1D–I; Supplementary Table S6). Accuracy was similar in the Glasgow and population-based FLEMENGHO cohorts (Pearson r 0 74 to 0 89; MAE 5 44 years in both), and was lower with greater disease burden (MAE 5 83 years with no major comorbidity, 8 71 with multiple; Supplementary Table S7).

Without kidney conditioning, the clock was biased by kidney function in the direction opposite to risk. Participants with macroalbuminuria appeared younger than those with normoalbuminuria (unconditioned age acceleration -1·75 against +3·56 years; conditioned +9 86 against +3 49 years), and the fifth of participants who appeared most biologically aged had the best-preserved filtration (Supplementary Table S7). The bias also weakened the mortality association: the age- and sex-adjusted HR per within-stratum SD of unconditioned age acceleration fell from 1 36 and 1 37 in the two highest eGFR categories to 0 97 in KDIGO G4–G5, and a flexible spline of measured eGFR in the outcome model did not restore it among the 1,905 participants with an eGFR below 60 mL/min/1 73 m (HR 1 05, 95% CI 0 93–1 20, against 1 22, 1 06–1 40, for k-UPBioAgeAcc). Conditioning applies no correction to the 3,649 participants in the KDIGO low-risk zone, assigned partly from peptide-estimated albuminuria (Supplementary Methods S4, S7; Supplementary Figure S4).

k-UPBioAgeAcc was higher with prevalent diabetes, cardiovascular disease, kidney disease, and hypertension (Figure 2A). Relative to participants with none of the four, it was 1 2 years higher (95% CI 0 3–2 1) with hypertension alone, 12 7 years higher (11 7–13 7) with both diabetes and kidney disease, and 15 6 years higher (13 0–18 1) in the 38 participants with all four. Kidney disease alone carried an excess of 9 7 years (8 8–10 7; n=434), part of which reflects the kidney conditioning itself. Across the KDIGO risk grid, k-UPBioAgeAcc generally rose as filtration fell and albuminuria increased, and was highest where the lowest eGFR met the highest albuminuria (Figure 2B).

**Figure 2.**
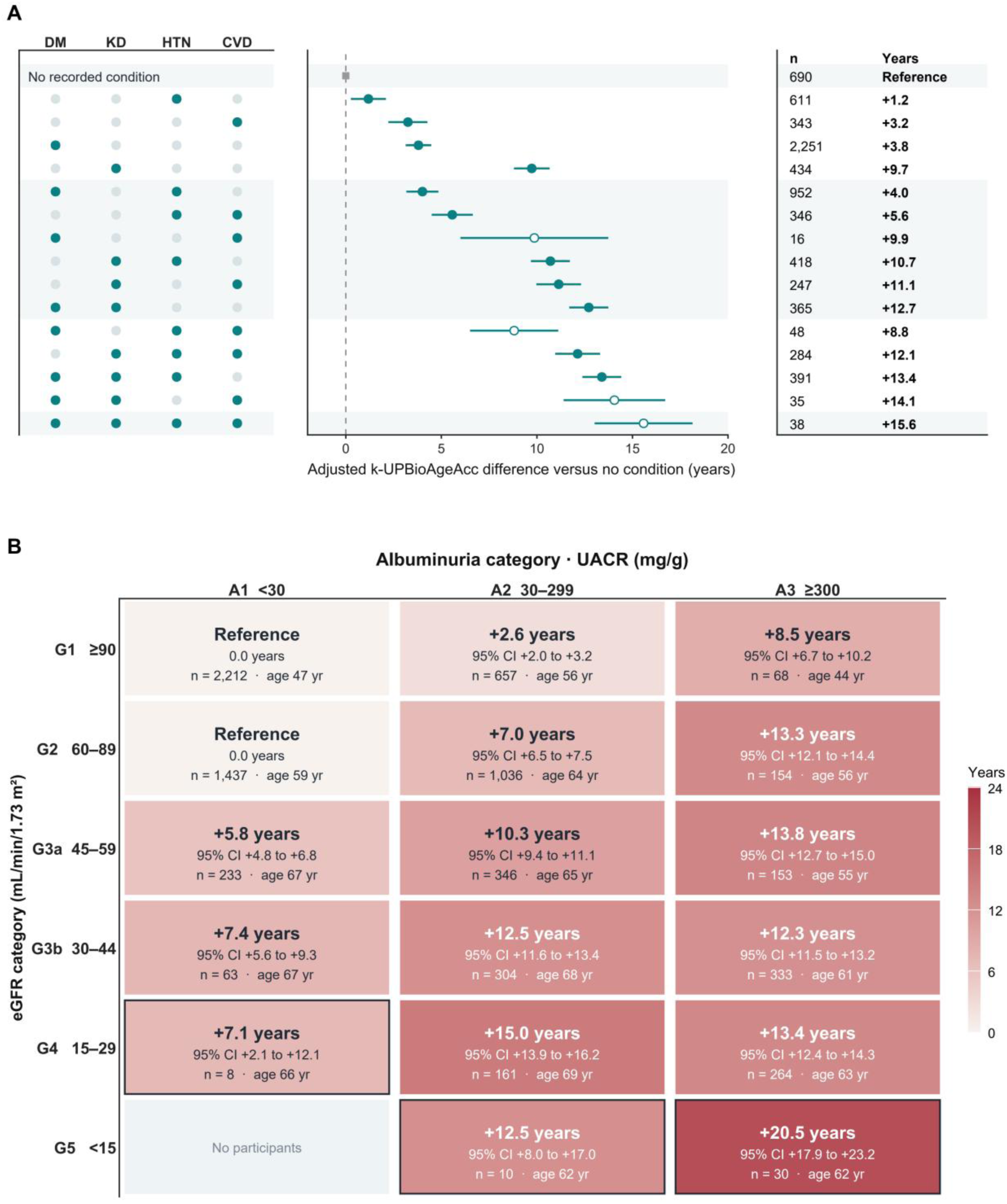
Kidney-conditioned urinary age acceleration across disease burden and the KDIGO risk grid. A, Condition combinations, each shown once; markers are point estimates with 95% CIs. Filled dots identify the conditions present: diabetes (DM), kidney disease (KD), hypertension (HTN), cardiovascular disease (CVD); pale dots indicate absence. Values are differences in k-UPBioAgeAcc from the no-condition reference (n=690); open markers indicate groups with fewer than 50 participants. B, Difference in k-UPBioAgeAcc from the pooled KDIGO low-risk zone (the pooled G1/A1 and G2/A1 cells, n=3,649, whose own mean k-UPBioAgeAcc is +1 37 years). Cells outlined in black contain fewer than 50 participants; a cell marked “No participants” indicates that no participant falls in that KDIGO combination. UACR uses measured values where available (2,482 of 7,469 participants) and peptide-based estimates otherwise, the latter for most participants in stages G3b–G5. Differences were estimated by linear regression, in panel A with adjustment for age, sex, body-mass index, and mean arterial pressure (the four conditions define the groups and are therefore not covariates), and in panel B with adjustment for age, sex, diabetes, cardiovascular disease, hypertension, body-mass index, and mean arterial pressure. A, albuminuria category; CI, confidence interval; G, glomerular filtration rate category; KDIGO, Kidney Disease: Improving Global Outcomes; UACR, urinary albumin-to-creatinine ratio; k-UPBioAgeAcc, kidney-conditioned urinary peptidomic biological age acceleration.

k-UPBioAgeAcc (SD 9 3 years) was associated with all-cause mortality in age- and sex-adjusted and fully adjusted models (HR per SD 1 62, 95% CI 1 49–1 76, and 1 47, 1 34– 1 62; both p<0 0001; Figure 3; Supplementary Table S8), similarly in women (1 50; 1 26– 1 79) and men (1 45; 1 30–1 63; interaction p=0 53). Associations were consistent across age groups and comorbidity strata (no comorbidity interaction survived BH correction), but varied across CKD stages (interaction p=0 031), with no association in KDIGO stage G4 or in measured macroalbuminuria (Supplementary Table S9). In separate models, adjustment for eGFR attenuated the association by 10 2% and for albuminuria by 37 8%, but it persisted with both in the model (HR 1 27, 95% CI 1 15–1 40; Supplementary Table S10). There was no evidence of non-proportional hazards (p=0 20), and the association was similar in a landmark analysis at the median follow-up (HR 1 45 before, 1 54 after) and after excluding deaths in the first 0 5, 1, or 2 years (HR 1 50 to 1 53).

**Figure 3.**
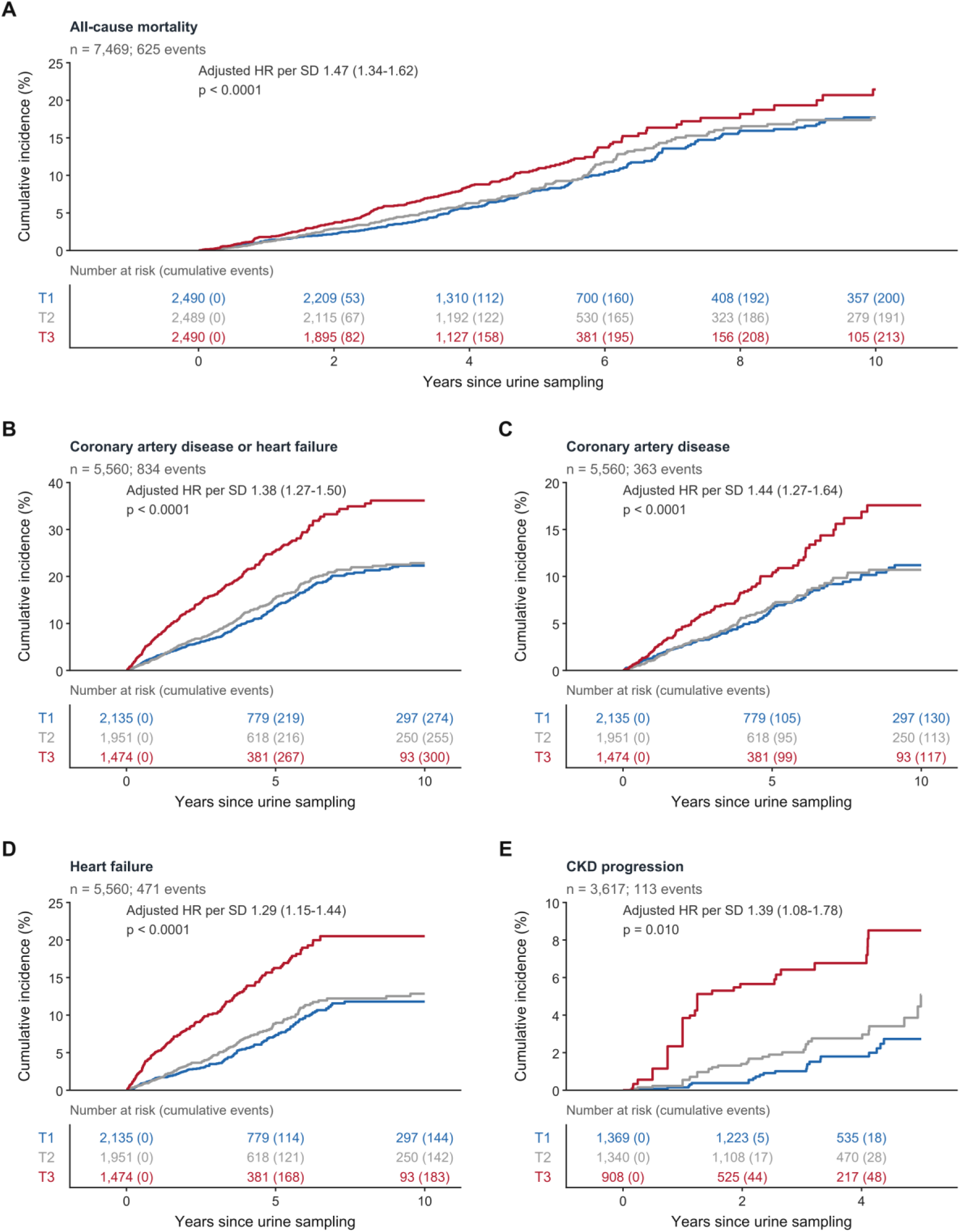
Cumulative incidence of death and incident disease by kidney-conditioned urinary age acceleration. Cumulative incidence by tertile of k-UPBioAgeAcc: T1, lowest (blue); T2, middle (grey); T3, highest (red); cut-points -0·04 and +7·54 years. A, All-cause mortality (n=7,469; 625 events). B, Coronary artery disease or heart failure (n=5,560; 834). C, Coronary artery disease (n=5,560; 363). D, Heart failure (n=5,560; 471). E, Chronic kidney disease progression (n=3,617; 113). Cumulative incidence is estimated by the Aalen– Johansen method, treating death before the endpoint as a competing event; 262, 151, and ten such deaths occurred in C, D, and E respectively (in E, deaths within kidney-function follow-up), and none in A or B. Curves are unadjusted; the hazard ratio printed in each panel is per 1 SD of k-UPBioAgeAcc from a cause-specific Cox model adjusted for age, sex, diabetes, cardiovascular disease, hypertension, body-mass index, mean arterial pressure, and eGFR. Numbers beneath each panel are participants at risk with cumulative events in parentheses, at the years marked on the axis. Panels A–D are truncated at 10 years, when 741 participants (9 9%) remain at risk for mortality, and panel E at 5 years, the last time at which all three tertiles are still observed; follow-up extends to 20 2 years. eGFR, estimated glomerular filtration rate; SD, standard deviation; k-UPBioAgeAcc, kidney-conditioned urinary peptidomic biological age acceleration.

The follow-up cohort combined 17 studies, of which 12 recorded enough deaths for a study-specific fully adjusted model (6,999 participants; 612 of the 625 deaths). Estimates were consistent across 11 of the 12 (random-effects pooled HR 1 38; 95% CI 1 19–1 59; I =39 8%; Supplementary Figure S5), and the association was attenuated but persisted when each study was given its own baseline hazard (HR 1 34; 1 22–1 48). Under full adjustment, the HR per SD was 1 47 for k-UPBioAgeAcc against 1 21 for unconditioned age acceleration, a ratio of 1 21 (95% CI 1 16–1 27); the unconditioned measure was stronger in none of 2,000 bootstrap resamples (Supplementary Table S11). By quintile, the most aged fifth of k-UPBioAgeAcc carried an age- and sex-adjusted HR of 3 66 (2 81–4 75) relative to the least aged, against 2 32 (1 75–3 08) for the unconditioned measure (Supplementary Figure S6).

Added to age and sex alone, k-UPBioAgeAcc raised Harrell’s C-index from 0 798 to 0 815 (ΔC +0·018, 95% CI 0·009–0 026); added to the fully adjusted model the gain was smaller, 0·830 to 0·835 (ΔC +0·005, 95% CI <0·001–0 009; Supplementary Table S12). Both gains were statistically significant. At five years, under full adjustment, the continuous net reclassification improvement was 0 20 (0 11–0 30) and the integrated discrimination improvement 0 017 (0 010–0 026).

Under the same fully adjusted model, k-UPBioAgeAcc was also associated with incident disease (Figure 3; Supplementary Table S8): the composite cardiovascular endpoint (834 events; HR per SD 1 38, 95% CI 1 27–1 50), CAD (363; 1 44, 1 27–1 64), and HF (471; 1 29, 1 15–1 44), each among the same 5,560 at risk (all p<0 0001), and CKD progression (113 events among 3,617; 1 39, 1 08–1 78; p=0 010). Treating death as a competing event gave a similar estimate for CKD progression (Fine–Gray subdistribution HR 1 37, 1 04– 1 81). All four associations remained significant after Holm correction, none showed evidence of non-proportional hazards, and cumulative incidence was highest in the top tertile for every endpoint (5-year incidence in the top tertile 10 2% for CAD, 16 3% for HF, and 8 5% for CKD progression; all log-rank p<0 0001). Unconditioned age acceleration, by contrast, was not associated with incident HF (1 08, 0 97–1 19) or CKD progression (1 03, 0 82–1 29) after full adjustment, and only weakly with CAD (1 13, 1 00–1 27) and the composite endpoint (1 11, 1 02–1 20; Supplementary Methods S6).

Applied to the same held-out participants with both models fixed in advance, k-UPBioAge estimated chronological age with lower error than the published UPP-age clock, which has no age-bias correction (MAE 4 94 against 9 45 years; 5 69 for UPBioAge before that correction), and its age acceleration was more strongly associated with mortality (fully adjusted HR per SD 1 47 against 1 24; Supplementary Methods S3; Supplementary Table S13).

## Discussion

Conditioned on kidney function, a urinary peptidomic ageing clock measured in a single urine sample identified people at higher risk of death and incident disease, beyond their chronological age and established clinical risk factors. Each SD of k-UPBioAgeAcc carried a 47% higher hazard of death and a 29–44% higher hazard of incident CAD, HF, and CKD progression, and k-UPBioAgeAcc was higher in people with more chronic conditions. Without conditioning, the same clock was more weakly associated with mortality (HR 1 21 against 1 47 per SD) and, after full adjustment, not with incident HF or CKD progression. Urine can report on biological ageing, but most informatively once the kidney that produces it is taken into account.

The clinical consequence is that an unconditioned urinary clock falters where risk is highest. Glomerular filtration, albumin permeability, and tubular handling shape which peptides reach the urine, and because preserved filtration also predicts survival, kidney-related variation in an unconditioned clock runs against its ageing signal: it rated people with the healthiest kidneys as the most aged and lost its prognostic value as kidney function declined, a decline common in older adults.^15,16^ Adding eGFR to the outcome model did not repair the unconditioned score, which suggests that the distortion lies in the measurement itself and is better corrected in the score than in the outcome model. Conversely, k-UPBioAgeAcc is not only a marker of impaired kidney function: its association with mortality persisted after adjustment for both eGFR and albuminuria. Conditioning does not remove kidney biology from the marker; it re-orients kidney-related variation so that it runs with mortality risk rather than against it, and leaves the prediction unchanged in almost half of the follow-up cohort.

The same issue may arise for other biofluids whose composition reflects the function of the organ that produces them, such as saliva or cerebrospinal fluid. We call the resulting principle biofluid gating: the function of the producing organ is treated as part of the assay and conditioned on before the measurement is read as systemic biology. This study tests the principle for urine and the kidney only (Supplementary Methods S8).

Collagen-derived peptides accounted for most of the age estimate, pointing to extracellular-matrix turnover. Fragments of the same collagen chain moved in opposite directions with age, a pattern that bulk changes in protein abundance cannot easily explain; it suggests age-related changes in proteolytic processing, consistent with work on matrix ageing and urinary collagen fragments.^2,19–21^ Beyond the matrix, the retained peptides trace vascular, inflammatory, epithelial, and renal processes, although CE–MS cannot establish their tissue of origin. They include a fragment compatible with the endothelial marker PECAM-1,^22^ and matrix Gla protein, an inhibitor of vascular calcification whose urinary concentration has been associated with all-cause and cardiovascular mortality in FLEMENGHO.^23^

The earlier 54-peptide UPP-age clock showed that urinary peptide profiles carry substantial information about chronological age.^11^ k-UPBioAge extends that work with a broader peptide panel, non-linear modelling, and conditioning on kidney function, and in the same participants it outperformed UPP-age in both age prediction and mortality association. Its added discrimination beyond established risk factors was small but statistically significant, as for epigenetic and plasma proteomic clocks, whose strong mortality associations likewise yield modest gains in C-index.^3,5,24^

An ageing biomarker is also expected to respond to intervention, and urinary peptidomics offers a route to testing this. Disease-specific urinary classifiers, including CKD273, HF2, and CAD160, have been validated in independent cohorts,^18,25,26^ treatment-induced peptide changes have been carried through them,^27^ and recent work predicts which interventions might most favourably change an individual profile,^28^ although none of this yet shows that treatments so selected improve outcomes. UPBioAge shares this architecture: Figure 4 carries peptide changes reported in published intervention studies through the model to show how candidate interventions could be prioritised for trials. These are projections, not observed treatment responses (Supplementary Methods S9); we will test them against measured treatment responses in a dedicated follow-up study. Current frameworks ask ageing biomarkers for analytical validity, association with future outcomes, and responsiveness to intervention.^29^ This study provides evidence for predictive performance, transferability across cohorts, and association with prospectively recorded outcomes. Short-term within-person variation was low for the earlier UPP-age clock (mean coefficient of variation 7 3% across six samples over one month in three healthy individuals),^11^ but repeatability remains to be confirmed for UPBioAge, as does responsiveness to intervention. The urinary peptidome does, however, respond to external exposures: it changes with pharmacological and lifestyle interventions, and UPP-age is higher with greater long-term air-pollution exposure.^30^

**Figure 4.**
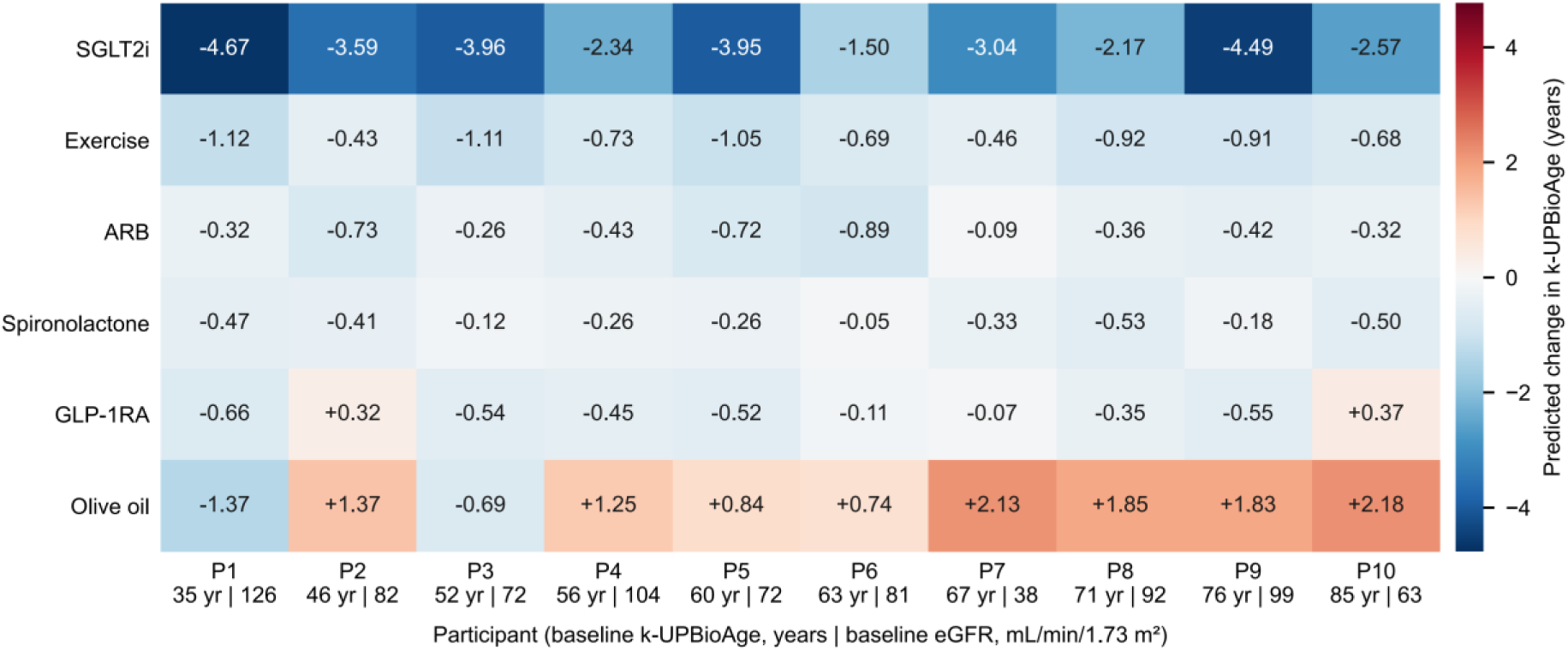
Predicted change in kidney-conditioned urinary peptidomic biological age across interventions. Predicted change in k-UPBioAge. (years) for six interventions in ten participants spanning the range of baseline k-UPBioAge. Blue, a predicted decrease; red, a predicted increase. Columns are individual participants, labelled P1 to P10, with their baseline k-UPBioAge (years) and baseline eGFR (mL/min/1 73 m ) printed beneath; rows are the evaluated interventions. These are model projections, not observed treatment responses, and carry no causal interpretation. Participants were selected deterministically as those nearest the 5th to 95th percentiles of baseline k-UPBioAge in ten-percentile steps across all 7,469 participants; no selection was based on predicted response. ARB, angiotensin-receptor blocker; GLP-1RA, glucagon-like peptide-1 receptor agonist; SGLT2i, sodium–glucose cotransporter-2 inhibitor; eGFR, estimated glomerular filtration rate; k-UPBioAge, kidney-conditioned urinary peptidomic biological age.

The k-UPBioAge clock was evaluated in largely independent populations differing in age, kidney function, and clinical characteristics, and every participant in the follow-up cohort had a measured eGFR. Several limitations apply. Recruitment was predominantly through European studies, but ethnicity and ancestry were not recorded, so ancestry-specific generalisability cannot be evaluated; the Glasgow cohort, though largely South African, contributes age prediction only. The follow-up cohort was clinically enriched, so the findings speak to relative rather than absolute risk, and the outcome associations need validation in general-population and non-European cohorts. The conditioned clock was not associated with mortality in KDIGO stage G4, possibly because uraemia and filtration failure outweigh the ageing signal there. Because kidney disease is unevenly distributed by ethnicity and socioeconomic position, a clock conditioned on kidney function inherits inequities in its measurement, including the race coefficient once used in creatinine-based eGFR. Measured albuminuria was available for 2,482 of 7,469 participants and estimated from the peptide data in the remainder, and imprecision in those estimates propagates into k-UPBioAgeAcc. The analyses are retrospective, endpoint ascertainment was not identical across studies, and cause of death was unavailable, so cause-specific mortality could not be examined. Medication was not recorded, so treatment effects on urinary peptides and prognosis may cause residual confounding.

Conditioned on kidney function, k-UPBioAgeAcc captured an ageing-related signal associated with mortality, incident disease, and multimorbidity. Measured in a sample collected without a needle, it could help identify older and multimorbid patients whose risk exceeds what their age and clinical profile suggest, and serve as a response measure in trials of interventions targeting ageing. Before such use, it requires prospective validation in general-population and non-European cohorts, calibration of absolute risk, assessment of within-person repeatability and responsiveness to intervention, and evidence that acting on it improves outcomes; until then, it is a research tool and not a basis for decisions about individual patients.

## Data Availability

The urinary peptidomic data are proprietary to Mosaiques Diagnostics GmbH and are available subject to data-use agreements. Anonymised summary statistics, and the coefficients, model weights and parent-protein annotations for all 2,631 peptides retained in the model (Supplementary Data 1), are provided as supplementary material to support independent evaluation and re-use. Individual-level data access can be requested from the corresponding author (H.M.).

## Contributors

SB developed the computational pipeline, performed all analyses, and wrote the manuscript. MAJC contributed to data processing and quality control. JS and AL contributed to peptidomic data generation and peptide sequencing. HM supervised the study and contributed to the study design and interpretation. MB contributed to clinical interpretation and supervised from the nephrology perspective. DSM contributed to the study design, statistical methodology, and interpretation, and critically revised the manuscript. TSN and JAS contributed to cohort data acquisition and curation and critically revised the manuscript. SB, HM, and DSM accessed and verified the underlying data. DSM and MB contributed equally. All authors had full access to all the data in the study, reviewed and approved the final manuscript, and had final responsibility for the decision to submit for publication.

## Declaration of interests

HM is the founder and co-owner of Mosaiques Diagnostics GmbH, which develops urinary proteomic diagnostics and CE–MS-based biomarker assays. SB, MAJC, JS, and AL are employees of Mosaiques Diagnostics GmbH. DSM, TSN, JAS, and MB declare no competing interests.

## Data sharing

Individual, de-identified participant data used in these analyses (urinary peptide intensities, age, sex, and the clinical and outcome variables analysed), together with a data dictionary, will be shared on request with any qualified investigator after approval of a protocol and signature of a data access agreement, for the analyses specified in the approved protocol, from publication with no end date. The urinary peptidomic data are proprietary to Mosaiques Diagnostics GmbH, and the approval of the investigators of the contributing studies may also be required. Requests should be sent to the corresponding author (HM). No study protocol or statistical analysis plan was prepared and the study was not registered, because this is a secondary analysis of existing anonymised data from studies that had already completed recruitment and follow-up; the informed consent forms are held by the contributing studies and will not be shared. The share of model weight and parent-protein annotation of each of the 2,631 peptides retained in the model are provided in Supplementary Data 1 to support independent evaluation and re-use.

Code to reproduce the model fitting, calibration, and statistical analyses, together with the complete fitted model (all Stage-1 fits, preprocessing constants, the Stage-2 coefficients, the age-bias correction, and the calibration surface; participant-level data removed), is deposited on Zenodo (doi:10.5281/zenodo.22914811). Editors and reviewers are given access during peer review, and the deposit will be made public on publication. It includes an environment specification (Python 3.12.10 with scikit-learn 1.8.0, XGBoost 3.0.5, lifelines 0.30.1, statsmodels 0.14.5, and SciPy 1.15.3), the scripts that reproduce the supplementary analyses and Figures 1–3 and Supplementary Figures S1–S6 from a completed run, and the complete g:Profiler result table of the pathway analysis.

## Acknowledgements

We thank all participants and data contributors across all the cohorts used in this study. We acknowledge the FLEMENGHO study investigators for access to the FLEMENGHO validation cohort. There was no patient or public involvement in the design, conduct, reporting, or dissemination of the present analysis.

## Funding

This work received funding from the European Union’s Horizon Europe Marie Skłodowska-Curie Actions Doctoral Networks programme through the PICKED project (HORIZON-MSCA-2023-DN-01, Grant Agreement No. 101168626). This work was also supported in part by the German Federal Ministry of Education and Research (BMBF) through the ERA PerMed SIGNAL project (01KU2307), and by the COST Actions PerMediK (CA21165) and KidneyHealth (CA25128). Funded by the European Union. Views and opinions expressed are however those of the authors only and do not necessarily reflect those of the European Union or the European Research Executive Agency (REA). Neither the European Union nor the granting authority can be held responsible for them.

## Abbreviations

ARB: angiotensin-receptor blocker.
BH: Benjamini–Hochberg.
BMI: body-mass index.
CAD: coronary artery disease.
CE–MS: capillary electrophoresis–mass spectrometry.
CI: confidence interval.
CKD: chronic kidney disease.
CKD-EPI: Chronic Kidney Disease Epidemiology Collaboration.
COVID-19: coronavirus disease 2019.
CVD: cardiovascular disease.
DM: diabetes.
eGFR: estimated glomerular filtration rate.
FAC: Filtrate-Aware Calibration.
FLEMENGHO: Flemish Study on Environment, Genes and Health Outcomes.
GLP-1RA: glucagon-like peptide-1 receptor agonist.
HF: heart failure.
HR: hazard ratio.
HTN: hypertension.
KD: kidney disease.
KDIGO: Kidney Disease: Improving Global Outcomes.
k-UPBioAge: kidney-conditioned urinary peptidomic biological age.
k-UPBioAgeAcc: kidney-conditioned urinary peptidomic biological age acceleration.
LC–MS/MS: liquid chromatography–tandem mass spectrometry.
MAE: mean absolute error.
PECAM-1: platelet endothelial cell adhesion molecule-1.
RMSE: root mean squared error.
SD: standard deviation.
SGLT2i: sodium–glucose cotransporter-2 inhibitor.
UACR: urinary albumin-to-creatinine ratio.
UPBioAge: urinary peptidomic biological age.
UPP-age: urinary peptidomic profiling age.

## Notes

### Competing Interest Statement

H.M. is the founder and co-owner of Mosaiques Diagnostics GmbH which develops urinary proteomic diagnostics and CE-MS-based biomarker assays. S.B., M.A.J.C., J.S. and A.L. are employees of Mosaiques Diagnostics GmbH. D.S.M., T.S.N., J.A.S. and M.B. declare no competing interests.

### Author Declarations

All datasets were from previously published studies and fully anonymised; ethical review and approval were waived for the present analysis by the ethics committee of Hannover Medical School, Germany (No. 3116-2016), due to all data being fully anonymised.

### Summary of Updates

This version was revised for journal submission. Main changes: Final analysis run. All results were recomputed from a locked run that (i) caps measured eGFR at 150 mL/min per 1.73 m2 when fitting the kidney-conditioning (Filtrate-Aware Calibration) surface, (ii) uses a re-selected peptide-based eGFR surrogate for participants without measured eGFR, and (iii) corrects one missing chronic kidney disease follow-up time. Estimates changed slightly and the conclusions are unchanged: holdout mean absolute error 4.94 years (previously 4.91); fully adjusted hazard ratio per SD for all-cause mortality 1.47, 95 percent CI 1.34 to 1.62 (previously 1.48, 1.35 to 1.63); heart failure 1.29 (1.27); chronic kidney disease progression 1.39, 1.08 to 1.78 (1.35, 1.05 to 1.73). New analyses of why kidney conditioning is needed. Without conditioning, the mortality association of the clock weakened as eGFR fell and was lost in KDIGO stages G4 to G5, and adding a flexible spline of measured eGFR to the outcome model did not restore it in participants with eGFR below 60. We also added a model with a separate baseline hazard for each contributing study and Holm correction across the incident-disease endpoints. Structure and length. The manuscript was restructured into a structured Summary, a Research in context panel and a shorter main text. The title was changed. Former Tables 2 and 3 (top contributing peptides and age-prediction performance) are now Supplementary Tables S4 and S6; Table 1 remains in the paper. Interpretation. Claims were moderated throughout. The clock is presented as a research tool that requires prospective validation, and the in-silico intervention projections are described explicitly as model projections, not observed treatment responses. Limitations were expanded (clinically enriched cohort, ancestry not recorded, medication not recorded, estimated albuminuria in part of the cohort). Statements. The ethics statement now states that the Ethics Committee of Hannover Medical School has waived the requirement for ethical approval for analyses of fully anonymised data (No. 3116-2016), and that approval and written informed consent were obtained in each original study. The data sharing statement was rewritten, the analysis code and fitted model are deposited on Zenodo (doi:10.5281/zenodo.22914811; public on publication), the COST Action KidneyHealth (CA25128) was added to the funding statement, and STROBE and TRIPOD-AI reporting is stated. Supplementary Appendix. Updated to the final analysis run, with expanded methods for the kidney conditioning, the sensitivity analyses and the per-study composition of the follow-up cohort.

## References

1 Partridge L, Deelen J, Slagboom PE. Facing up to the global challenges of ageing. Nature 2018; 561: 45–56.

2 López-Otín C, Blasco MA, Partridge L, Serrano M, Kroemer G. Hallmarks of aging: An expanding universe. Cell 2023; 186: 243–78.

3 Moqri M, Herzog C, Poganik JR, et al. Biomarkers of aging for the identification and evaluation of longevity interventions. Cell 2023; 186: 3758–75.

4 Rutledge J, Oh H, Wyss-Coray T. Measuring biological age using omics data. Nat Rev Genet 2022; 23: 715–27.

5 Argentieri MA, Xiao S, Bennett D, et al. Proteomic aging clock predicts mortality and risk of common age-related diseases in diverse populations. Nat Med 2024; 30: 2450–60.

6 Mutz J, Iniesta R, Lewis CM. Metabolomic age (MileAge) predicts health and life span: A comparison of multiple machine learning algorithms. Sci Adv 2024; 10: eadp3743.

7 Latosinska A, Siwy J, Mischak H, Frantzi M. Peptidomics and proteomics based on CE-MS as a robust tool in clinical application: The past, the present, and the future. Electrophoresis 2019; 40: 2294–308.

8 Decramer S, de Peredo AG, Breuil B, et al. Urine in Clinical Proteomics. Mol Cell Proteomics 2008; 7: 1850–62.

9 Hepburn S, Cairns DA, Jackson D, et al. An analysis of the impact of pre-analytical factors on the urine proteome: Sample processing time, temperature, and proteolysis. Proteomics Clin Appl 2015; 9: 507–21.

10 Nkuipou-Kenfack E, Bhat A, Klein J, et al. Identification of ageing-associated naturally occurring peptides in human urine. Oncotarget 2015; 6: 34106–17.

11 Martens DS, Thijs L, Latosinska A, et al. Urinary peptidomic profiles to address age-related disabilities: a prospective population study. Lancet Healthy Longev 2021; 2: e690–703.

12 Lindeman RD, Tobin J, Shock NW. Longitudinal Studies on the Rate of Decline in Renal Function with Age. J Am Geriatr Soc 1985; 33: 278–85.

13 Magalhães P, Pontillo C, Pejchinovski M, et al. Comparison of Urine and Plasma Peptidome Indicates Selectivity in Renal Peptide Handling. Proteomics Clin Appl 2018; 12: e1700163.

14 Catanese L, Siwy J, Mischak H, Wendt R, Beige J, Rupprecht H. Recent Advances in Urinary Peptide and Proteomic Biomarkers in Chronic Kidney Disease: A Systematic Review. Int J Mol Sci 2023; 24: 9156.

15 Delanaye P, Jager KJ, Bökenkamp A, et al. CKD: A Call for an Age-Adapted Definition. J Am Soc Nephrol 2019; 30: 1785–805.

16 Stevens PE, Ahmed SB, Carrero JJ, et al. KDIGO 2024 Clinical Practice Guideline for the Evaluation and Management of Chronic Kidney Disease. Kidney Int 2024; 105: S117–314.

17 Keller F, Beige J, Siwy J, et al. Urinary peptides provide information about the risk of mortality across a spectrum of diseases and scenarios. J Transl Med 2023; 21: 663.

18 Good DM, Zürbig P, Argilés À, et al. Naturally Occurring Human Urinary Peptides for Use in Diagnosis of Chronic Kidney Disease. Mol Cell Proteomics 2010; 9: 2424–37.

19 Martin EM, Genovese F, Mischak H, et al. Association of Urinary Collagen Type III Degradation Product With Kidney Function and Fibrosis in Chronic Kidney Disease Patients. Proteomics 2025; 25: e202400354.

20 Statzer C, Park JYC, Ewald CY. Extracellular Matrix Dynamics as an Emerging yet Understudied Hallmark of Aging and Longevity. Aging Dis 2023; 14: 670–93.

21 Preston R, Hoyle A, Stevenson Harris A, et al. Ageing impacts extracellular matrix turnover and remodelling in the kidney. Matrix Biol Plus 2026; 30: 100197.

22 Caligiuri G. CD31 as a Therapeutic Target in Atherosclerosis. Circ Res 2020; 126: 1178–89.

23 Wei D, Melgarejo J, Vanassche T, et al. Urinary matrix Gla protein is associated with mortality risk in Flemish population: A prospective study. Front Cardiovasc Med 2022; 9: 894447.

24 Xiao H, Lau C-HE, Dehghan A, Robinson O. Proteomic aging clocks in epidemiological studies: advances, applications and prospects. Nat Aging 2026; 6: 970–86.

25 Zhang Z, Staessen JA, Thijs L, et al. Left ventricular diastolic function in relation to the urinary proteome: A proof-of-concept study in a general population. Int J Cardiol 2014; 176: 158–65.

26 Wei D, Melgarejo JD, Van Aelst L, et al. Prediction of coronary artery disease using urinary proteomics. Eur J Prev Cardiol 2023; 30: 1537–46.

27 Jaimes Campos MA, Andújar I, Keller F, et al. Prognosis and Personalized In Silico Prediction of Treatment Efficacy in Cardiovascular and Chronic Kidney Disease: A Proof-of-Concept Study. Pharmaceuticals 2023; 16: 1298.

28 Latosinska A, Mina IK, Nguyen TMN, et al. In silico prediction of optimal multifactorial intervention in chronic kidney disease. J Transl Med 2025; 23: 943.

29 Moqri M, Herzog C, Poganik JR, et al. Validation of biomarkers of aging. Nat Med 2024; 30: 360–72.

30 Martens DS, An D-W, Yu Y-L, et al. Association of Air Pollution with a Urinary Biomarker of Biological Aging and Effect Modification by Vitamin K in the FLEMENGHO Prospective Population Study. Environ Health Perspect 2023; 131: 127011.

