## Supplemental Material for "A kidney-conditioned urinary peptidomic biological ageing clock and its association with all-cause mortality and age-related health outcomes: a retrospective multicohort development and validation study"

**Contents** Page


**Supplementary Methods**

S1 Study cohorts and eligibility ... 3
S2 Urinary peptidomics by CE–MS ... 4
S3 Two-stage clock model, post-processing, UPBioAge, and UPBioAgeAcc generation ... 4
S4 Kidney surrogates and Filtrate-Aware Calibration ... 7
S5 Mortality analysis ... 10
S6 Incident events and disease-burden age acceleration ... 11
S7 Sensitivity and robustness analyses ... 12
S8 Enrichment, statistics, reproducibility, and supporting detail ... 15
S9 In-silico intervention analysis ... 17

**Supplementary Figures**

S1 Study cohorts and analytical workflow ... 19
S2 Per-peptide age-correlation volcano ... 20
S3 Pathway over-representation of age-clock peptide proteins ... 21
S4 Kidney-surrogate model accuracy ... 22
S5 Per-study meta-analysis of the association of UPBioAgeAcc and k-UPBioAgeAcc with all-cause mortality ... 23
S6 Filtrate-Aware Calibration reverses a biologically implausible ordering ... 24

**Supplementary Tables**

S1 Per-study composition of the clinical follow-up cohort ... 25
S2 Incidence, follow-up, and number of adverse health outcomes in the clinical follow-up cohort ... 26
S3 Clinical follow-up cohort demographic characteristics ... 27
S4 Top 20 contributing peptides to UPBioAge ... 28
S5 Pathway over-representation of the age-clock peptide proteins ... 28
S6 Chronological-age prediction performance of the k-UPBioAge clock across analysis cohorts ... 31
S7 Prediction accuracy of k-UPBioAge, and the effect of kidney conditioning on age acceleration, by cohort and by clinical stratum ... 32
S8 Risk for adverse outcomes in association with kidney-conditioned urinary peptidomic age acceleration (k-UPBioAgeAcc) ... 33
S9 Mortality association of k-UPBioAgeAcc within strata of the clinical follow-up cohort, by CKD stage, albuminuria category, age group, sex, and comorbidity ... 34
S10 eGFR and UACR attenuation of the k-UPBioAgeAcc mortality association ... 36
S11 Mortality and incident-disease associations of the kidney-conditioned and unconditioned urinary age acceleration, side by side ... 37
S12 Discrimination for all-cause mortality, with and without the kidney-conditioned urinary age acceleration ... 39
S13 Head-to-head comparison against the published UPP-age clock ... 40
S14 Mortality association of k-UPBioAgeAcc according to whether a Filtrate-Aware correction was applied, and according to whether albuminuria was measured or estimated ... 42

**Supplementary Data**

1 Peptide-level model detail ........ 43

**Supplementary Methods S1: Study cohorts and eligibility**

**Independence of the cohorts.** All validation cohorts were independent of model development: no validation participant contributed to clock derivation or to the Filtrate-Aware Calibration (FAC) modelling. The validation cohorts are not, however, disjoint from one another, because 775 participants of the Flemish Study on Environment, Genes and Health Outcomes (FLEMENGHO) also belong to the clinical follow-up cohort. They serve two separate purposes: FLEMENGHO validates the age predictions in a population cohort, and its 775 shared participants contribute, with the rest of the clinical follow-up cohort, to the outcome analyses.

**Sample collection dates.** Urine was collected between 1985 and 2024 in the derivation cohort (95% of dated samples 2002–2019; date recorded for 86·9%), between 1985 and 2022 in the kidney-diverse cohort (2002–2021; 79·8%), between 2002 and 2016 in the Glasgow samples, between 2005 and 2011 in FLEMENGHO (dates recorded for all participants in both), and between 1999 and 2019 in the clinical follow-up cohort (2002–2016; 98·9%; per study in Supplementary Table S1). Dates are those recorded for each sample in the laboratory sample register.

**Earlier analyses of the clinical follow-up cohort.** The mortality data of the clinical follow-up cohort have been analysed in two earlier publications from our group, both using classifiers trained on clinical outcomes rather than on age: the previously developed COV50 score, applied to mortality risk across a spectrum of diseases,[1] and a collagen-peptide mortality classifier (COL210), developed in a separate COVID-19 cohort and validated in 7,474 participants of the same cohort at an earlier data freeze.[2] The present clock differs from both: it was trained on chronological age in a separate derivation cohort, no outcome information was used in developing the clock or the calibration, and neither earlier analysis conditions on kidney function.

**Derivation cohort eligibility.** The albuminuria threshold was held constant at all ages because it marks glomerular-barrier damage specifically, whereas the filtration threshold was age-adapted because age-related decline in estimated glomerular filtration rate (eGFR) is largely physiological and a fixed cut-off over-diagnoses chronic kidney disease (CKD) in older adults. Participants were recorded as healthy controls in the source database, with no documented history of the excluded conditions. No eGFR was recalculated for this work: every eGFR value was taken as reported by the contributing study, each of which used a creatinine-based Chronic Kidney Disease Epidemiology Collaboration (CKD-EPI) equation.

**FLEMENGHO.** FLEMENGHO is a prospective family-based population study that recruited a random sample of households from a geographically defined area of northern Belgium and has followed them with repeated standardised examinations, providing a general-population comparator to the clinically enriched follow-up cohort.

**Clinical follow-up cohort**. Included studies were EU-PRIORITY, DIRECT, EPOGH, Sun-MACRO, UZ Gent, Generation Scotland, uPROPHET, HOMAGE fibrosis, PersTIgAN, CAD Predictions, TransBioBC, CardioRen, STOP-IgAN, Heart failure and Predictions (HIB, Prague, and Groningen) with detailed information provided in Supplementary Table S1. Diagnoses were available for diabetes (type 1 or type 2), kidney disease (CKD, IgA nephropathy, diabetic nephropathy, membranous nephropathy, minimal change disease, lupus nephritis, focal segmental glomerulosclerosis), cardiovascular disease (coronary artery disease [CAD], heart failure [HF], heart transplantation) and hypertension. Hypertension was defined as a blood pressure of at least 140 mmHg systolic or at least 90 mmHg diastolic. Urine was collected between May 1999 and April 2019 and participants were followed to a maximum of November 2020.

**Kidney-diverse cohort for surrogate modelling and FAC. No kidney-function criterion was applied**

when selecting this cohort, so participants with diagnosed kidney disease were included alongside those with preserved filtration; they were selected to enable the inclusion of a wide range of eGFR and urinary albumin-to-creatinine ratio (UACR) values, resulting in a cohort of 7,797 participants. Of these, 7,540 had measured eGFR (CKD-EPI equation) and 4,946 had measured UACR.

**Supplementary Methods S2: Urinary peptidomics by CE–MS**

Urine samples (midstream second-void collections) were processed by capillary electrophoresis–mass spectrometry (CE–MS) as previously described.[3] Samples were thawed and diluted 1:1 (v/v) with buffer including 2 M urea, 10 mM $NH_4OH$, and 0·02% SDS, ultrafiltered through a 20 kDa molecular-weight cut-off centrifugal device (Centrisart, Sartorius), desalted on PD-10 columns (GE Healthcare), lyophilised and reconstituted in 10 µL HPLC-grade water. CE separation used a 90 cm × 50 µm i.d. bare fused-silica capillary with 20% acetonitrile and 1% formic acid as background electrolyte (25 kV; 35 °C; hydrodynamic injection 2 psi, 99 s), coupled via a sheath-flow interface (50% isopropanol, 0·1% formic acid, 200 nL/min) to a Bruker micrOTOF II electrospray-ionisation time-of-flight (ESI-TOF) mass spectrometer in positive-ion mode (m/z 350–3,000, spectra every 3 s). Raw spectra were processed with MosaFinder for signal detection, charge-state deconvolution, and internal calibration against reference standards; normalised signal intensities served as relative abundances.[4] The date of each CE–MS measurement is recorded in its run identifier. Where it could be recovered (every derivation and FLEMENGHO sample, and 99·0% of the clinical follow-up and Glasgow samples), samples were measured between 2004 and 2024: the clinical follow-up cohort in 2007–2019, FLEMENGHO in 2010–2014, the Glasgow cohort in 2008–2018 (mostly 2016–2018), and the derivation cohort in 2004–2024. The median interval between collection and measurement was 3 years in the derivation, Glasgow, and clinical follow-up cohorts and 6 years in FLEMENGHO; it exceeded 10 years for 3·1% of the clinical follow-up cohort and for fewer than 2% of every other cohort. Sampling era and storage time were not adjusted for in the outcome models; in the clinical follow-up cohort, the year of collection correlated with age ($r = 0·30$). Peptide sequences were assigned in separate CE–MS/MS and liquid chromatography–tandem mass spectrometry (LC–MS/MS) runs. The two separations are used together because they identify substantially different and complementary parts of the urinary peptidome, so neither alone gives full sequence coverage.[5] Fragment spectra from both routes were searched with Proteome Discoverer 2·4 (Thermo Fisher Scientific) and SEQUEST against UniProt Swiss-Prot (canonical sequences; no enzyme specificity; precursor tolerance 5 ppm; fragment tolerance 0·05 Da; variable modifications oxidation of methionine and proline), retaining only high-confidence assignments (false discovery rate <1%). Sequences established in this way are held in the Human Urinary Proteome Database and are matched to the CE–MS signals of each new sample by mass and normalised migration time, so peptides are not re-sequenced sample by sample.

**Supplementary Methods S3: Two-stage clock model, post-processing, UPBioAge, and UPBioAgeAcc generation**

**Software, peptide screening and model overview.** All analyses were performed in Python 3.12.10 (scikit-learn 1.8.0, XGBoost 3.0.5, statsmodels 0.14.5, lifelines 0.30.1, SciPy 1.15.3). In a first step, peptides with a detection frequency of ≥ 40% across participants in the training set of the derivation cohort ($n = 1,449$) were selected (4,340 peptides). In a second step, these peptides were correlated with chronological age using Spearman rank correlation. Peptides that remained significant after correction for multiple-testing using the Benjamini–Hochberg (BH) procedure ($q < 0·05$) were selected for the urinary peptidomic biological age (UPBioAge) clock model development. In a third step, the UPBioAge clock was developed using a two-stage stacked-regression. In stage 1, chronological age was modelled independently on each retained peptide, as a quadratic function of that peptide's standardised abundance, producing individual peptide-based age predictions. The quadratic term captures the non-linear age trajectories as previously observed in plasma proteomic ageing studies.[6]

In stage 2, an elastic-net meta-model was fitted by cross-validation over a wide grid of l1-ratios and penalties using the individual peptide-based age predictions as input, retaining all peptides with a non-zero coefficient. The two-stage design follows the precedent established by the DNA methylation GrimAge clock.[7] The stage-1 predictions entering stage 2 were generated out of fold (OOF), as set out below.

**Stage 1** acts as a set of univariate, ridge-regularised, low-degree polynomial age models. For each of the 3,296 BH-significant age-correlated peptides, chronological age was regressed on the standardised abundance of that single peptide. Writing z for the abundance of one peptide, standardised to zero mean and unit variance in the training set, the fitted model is age $\approx \beta_0 + \beta_1 z + \beta_2 z^2$. The peptide is therefore the predictor and chronological age the response, and each peptide yields an age prediction per sample directly, with no inversion step. Coefficients were obtained by least squares on the two-degree polynomial basis (intercept, linear, quadratic), with a ridge term of $\lambda = 10^{-6}$ added only to condition the solution for sparsely detected peptides; at this magnitude it applies no material shrinkage. An undetected peptide is recorded as an intensity of zero, which the log(x+1) transformation keeps at zero, so it enters that peptide's fit and prediction as the lowest abundance rather than being excluded, and every sample receives a prediction from every peptide. A stage-2 median fill, computed once in the training set and applied unchanged to every held-out fold and validation cohort, is kept only as a safeguard for a prediction that cannot be formed; it was not needed for any sample in any cohort. Standardised abundances were not clipped. Peptide-level age predictions were bounded at 20 years below the youngest and 20 years above the oldest training participant (−2 to 106 years) before stage 2; the bound applied to fewer than 0·01% of peptide-level predictions in the derivation holdout and in every validation cohort. To prevent leakage into calibration, the stage-2 predictions used to fit the post-processing layers were generated from 5-fold OOF stage-1 predictions with folds stratified on age bins; the deployed stage-2 model was refitted on stage-1 predictions from models fitted to the full training set, with preprocessing fitted in that set. Full-training-set fits were used for holdout, external and mortality inference (deployment style). The deployed per-peptide age predictions may therefore have slightly lower variance than their OOF training counterparts; this is the expected distributional shift of leakage-safe stacked modelling and is monitored as a reproducibility diagnostic, not treated as target leakage.

**Stage 2** is an ensemble over the resulting per-peptide age predictions**.** An elastic-net meta-model (scikit-learn) was fitted on the 3,296 per-peptide UPBioAge predictions of the stage-1 models fitted to the full training set, with hyperparameters chosen by cross-validation over a grid of 41 l1-ratios × 200 alphas (8,200 candidate configurations; alpha $10^{-6}$–500), selecting the configuration with the minimum cross-validated mean squared error, which gave $\alpha = 0{\cdot}881$ and l1-ratio = 0·03 (coordinate descent with random coordinate selection, maximum 200,000 iterations, tolerance $1\times10^{-5}$, intercept fitted). The search used five internal folds (scikit-learn KFold, unshuffled, in training-set row order) on the training matrix z-scored once beforehand, the seed (42) fixing the coordinate-descent update order; it was run once with the archived selection script, and the deployed model was fitted with the selected values rather than re-searched. The final fitted model retained 2,631 of 3,296 peptides with a non-zero coefficient (each peptide's share of model weight in Supplementary Data 1; the complete fitted model, including every Stage-1 fit, the Stage-2 coefficients, the preprocessing constants, the spline correction, and the calibration surface, is in the Zenodo deposit, doi:10.5281/zenodo.22914811). Evaluation cohorts were scored by applying the previously fitted Stage-1 and Stage-2 models without further fitting. Because the 3,296-peptide BH screen was performed only within the training set (n = 1,449) and was not re-estimated inside each Stage-1 fold, derivation OOF metrics are reported as internal calibration metrics, and the independent holdout and external-validation performance constitute the primary prediction-accuracy claim.

**Post-processing.** After Stage 2, two sequential post-processing layers, fitted on training OOF predictions only, convert raw predictions to the final UPBioAge clock. First, a linear recalibration regressed chronological age on UPBioAge ($y_{true} \approx a + b\cdot\hat{y}_{raw}$), correcting mean bias and slope shrinkage introduced by high-dimensional regularised regression. Second, a regression-to-the-mean (RTM) spline correction (six-knot cubic B-spline basis, degree 3, uniform knots, constant extrapolation beyond the fitted range, Ridge α = 0·5, uniform weights) was fitted to training OOF residuals on chronological age over the 20–80 yr window and subtracted, yielding UPBioAge. Participants younger than 20 or older than 80 yr were excluded from the spline fit because the sparse age tails distort the correction (n = 18 and n = 8 excluded, respectively, leaving 1,423 of 1,449). The linear recalibration is a fixed function of the raw prediction, whereas the spline correction is a function of chronological age and is evaluated at the participant's own chronological age whenever the clock is applied, in the derivation set and in every validation cohort alike. Every accuracy figure reported for UPBioAge and its kidney-conditioned form (k-UPBioAge) is therefore a calibrated rather than an age-blind prediction error, as is standard for age-bias-corrected brain-age and epigenetic clocks. The blind prediction, taken after linear recalibration but before the age-dependent spline correction, gives a mean absolute error (MAE) of 5·69 yr (r = 0·913, $R^2$ = 0·831) in the internal holdout, 7·03 yr (r = 0·823) in FLEMENGHO, 8·97 yr (r = 0·606) in the Glasgow cohort and 8·45 yr (r = 0·718) in the clinical follow-up cohort, against 4·87, 5·41, 5·41, and 6·78 yr respectively after the spline correction (4·94, 5·44, 5·44, and 7·93 yr for k-UPBioAge). The two age acceleration measures that follow from these two predictions are highly correlated, with an r of 0·988 in the derivation training set and r = 0·995 in the clinical follow-up cohort, with mean absolute differences of 1·91 and 2·10 yr respectively (mean signed differences 0·00 and 0·11 yr). The choice makes no material difference to any reported association: the fully adjusted mortality hazard ratio (HR) per standard deviation (SD) for the unconditioned UPBioAgeAcc is 1·213 with the spline correction applied and 1·213 without it. That comparison is between two versions of the same unconditioned measure and is separate from the unconditioned-versus-kidney-conditioned comparison of Supplementary Table S11. Post-hoc correction is important because residual bias propagates directly into age acceleration estimates, as shown for epigenetic clocks;[8] regression-to-the-mean bias is neither data- nor method-specific across algorithms,[9] and the CE–MS platform has documented analytical repeatability for urinary peptides and established classifier scores in repeated measurements of pooled urine,[3] and the earlier UPP-age clock on the same platform varied within individuals by a mean coefficient of variation of 7·3% (range 2·7–11·3%) across six urine samples collected within one month in three healthy individuals,[10] although the repeatability of UPBioAge itself within individuals remains to be established. Both age-acceleration measures (UPBioAgeAcc and k-UPBioAgeAcc, the residuals of UPBioAge and k-UPBioAge on chronological age) are orthogonal to chronological age by construction, and in the derivation OOF set the observed correlation is r < 0·001 for both, because UPBioAgeAcc is the residual from regressing UPBioAge on chronological age in the training-OOF set, consistent with published best-practice guidance for multi-organ ageing clocks.[11]

k-UPBioAgeAcc is defined the same way, by regressing k-UPBioAge on chronological age in the training OOF set. For both measures the intercept and slope were estimated once in that set and then held fixed; neither was re-estimated in any validation or outcome cohort. The two acceleration measures are therefore orthogonal to chronological age by construction only within the derivation OOF set, and retain a residual correlation with age elsewhere (−0·47 for UPBioAgeAcc and −0·33 for k-UPBioAgeAcc in the clinical follow-up cohort), which is the expected consequence of freezing the coefficients rather than refitting them within each cohort. Writing y for chronological age and $\beta_0$, $\beta_1$ for the intercept and slope of that fixed regression (and $\beta_0'$, $\beta_1'$ for the corresponding k-UPBioAge regression):

$$UPBioAgeAcc = \hat{y}UPBioAge - (\beta_0 + \beta_1 y) \quad and \quad k\text{-}UPBioAgeAcc = \hat{y}k\text{-}UPBioAge - (\beta_0' + \beta_1' y) \ (1)$$

**Head-to-head comparison.** The published urinary peptidomic profiling age (UPP-age) predictor (54 peptides from 17 parent proteins) was applied to the same samples using its published coefficients and peptide bounds, without re-fitting, so that the two clocks were evaluated in identical participants. Its age predictions were scored as published, without an age-bias correction; for the mortality analyses its acceleration was formed, as for UPBioAge, by a linear and a spline residualisation on chronological age fitted in the 1,449 training participants. Age-prediction accuracy was compared using the same metrics as elsewhere (MAE, root mean squared error [RMSE], Pearson r, and $R^2$), and the association of each clock's age acceleration with all-cause mortality was estimated per SD of that measure in the clinical follow-up cohort under the same age- and sex-adjusted and fully adjusted models. Results are reported in Supplementary Table S13.

**Supplementary Methods S4: Kidney surrogates and Filtrate-Aware Calibration**

**Kidney surrogate models.** Urinary peptide-based surrogate models for eGFR and UACR were trained using XGBoost in the kidney-diverse cohort. Their predictions were then recalibrated for regression-to-the-mean compression with a cubic spline-Ridge map (three knots, Ridge $\alpha = 1{\cdot}0$) fitted against the measured values in the same cohort; eGFR values above 200 mL/min/1·73 m² (102 observations) were treated as missing when fitting that map. Creatinine-based eGFR was taken as reported by each contributing study, all of which used a creatinine-based CKD-EPI equation; no eGFR was recalculated here, ethnicity was not recorded, and no race coefficient was applied by the present investigators. Serum creatinine was available for only part of each cohort (46% of the kidney-diverse cohort), so eGFR was not recomputed with the 2021 race-free equation. eGFR had a mean of 78·8 mL/min/1·73 m² (SD 46·6; range 0·98–1,328; n = 7,540), whereas measured UACR had a mean of 772·3 mg/g (SD 1,741·2; range 0–39,411; n = 4,946).

**Handling of extreme kidney-function values.** Extreme upper values were capped before model training. Creatinine-based eGFR becomes unreliable at very low serum creatinine concentrations, including in individuals with reduced muscle mass or cachexia, and may overestimate filtration in young adults. Values above approximately 150 mL/min/1·73 m² also lie at or beyond commonly used thresholds for glomerular hyperfiltration and should not be interpreted as progressively better kidney function. eGFR was therefore capped at 150 mL/min/1·73 m². This affected 408 of 7,540 observations (5·4%): 306 were between 150 and 200 mL/min/1·73 m² and only four exceeded 500 mL/min/1·73 m². Including one observation already equal to 150, 409 observations lay at the upper boundary after capping.

UACR was capped at 5,000 mg/g. At very high values, the ratio becomes increasingly sensitive to low urinary creatinine, while values of this magnitude already indicate severe, nephrotic-range glomerular barrier dysfunction and provide limited additional quantitative discrimination. The cap affected 285 of 4,946 observations (5·8%), of which only four exceeded 10,000 mg/g.

Capping rather than exclusion retained participants with the most extreme kidney dysfunction while preventing implausible or unstable values from dominating model fitting. The same limits were applied to training targets, downstream surrogate predictions, and the measured eGFR used to fit the FAC surfaces.

UACR was transformed as ln(1 + UACR [mg/g]) before training, whereas eGFR was modelled on its original scale. The 3,296 age-correlated urinary peptides identified in the derivation cohort were used as candidate predictors for both surrogate models.

**Model tuning and validation.** Hyperparameters were selected using a deterministic, space-filling search with fold-specific early stopping. Generalisation performance of the complete tuning procedure was estimated by nested cross-validation, separately for eGFR (n = 7,540) and log-transformed UACR (n = 4,946), using five outer folds, three inner folds, and a prespecified search of 40 hyperparameter configurations. Within each outer training partition, the configuration with the lowest mean inner-fold RMSE was selected and evaluated only in the untouched outer partition. The accuracy shown in Supplementary Figure S4 is the out-of-fold performance of the selected configuration under five-fold cross-validation, not the nested estimate.

For the final deployable surrogate used in downstream analyses, one configuration per outcome was selected according to pooled inner-validation RMSE. The accuracy we report for the surrogates is OOF. The peptide-based eGFR and UACR values used to fit the estimate-based calibration surface are, however, predictions made by that final surrogate in the same kidney-diverse cohort that trained it, so for that one purpose they are in-sample. This applies fully to the estimate-based surface, which is used when a measured eGFR is missing and scores no participant in the clinical follow-up cohort, where every participant has a measured eGFR. It applies in part to the measured-coordinate surface, which used peptide-based albuminuria estimates for the 2,828 participants without a measured UACR when it was fitted (see Dual-arm calibration procedure). The age predictions both surfaces are fitted against are out of sample for the clock in every case. The number of boosting rounds was set to the median of its fold-specific early-stopping optima, with early stopping on a further 15% of each inner-training split; the deployed configurations were fixed from that search and not re-searched in the analysis run. Fixed hyperparameters are reported in the deposited model metadata, and surrogate performance is shown in Supplementary Figure S4. Because the surrogates are used to place participants in Kidney Disease: Improving Global Outcomes (KDIGO) categories as well as to supply continuous coordinates, their behaviour at the category boundaries was examined OOF in the kidney-diverse cohort. For albuminuria (n = 4,946) the surrogate reproduced the exact three-level KDIGO category in 75·2% of participants; it identified albuminuria of at least 30 mg/g with a sensitivity of 89·4% and a specificity of 76·6%, and albuminuria of at least 300 mg/g with a sensitivity of 84·7% and a specificity of 92·8%. For filtration (n = 7,540) the surrogate identified an eGFR below 60 mL/min/1·73 m² with a sensitivity of 67·4% and a specificity of 94·1%, and below 30 mL/min/1·73 m² with a sensitivity of 54·7% and a specificity of 98·9%. Both surrogates therefore regress towards the middle of their range, which is why measured values are used wherever they exist and why categories built on a resolved coordinate are not interchangeable with categories built on a measured one.

Following incorporation of the kidney surrogates into FAC, the residual regression-to-the-mean slope in the holdout cohort was +0·016, against +0·003 after spline correction alone; both are close to zero.

**FAC. Let y** denote chronological age and $\hat{y}lin$ the linearly recalibrated prediction, taken before the age-dependent spline correction; the fitted correction is added to the spline-corrected UPBioAge (equation 4). Prediction error was assumed to vary systematically with kidney function, because kidney function influences the urinary peptidome, and was defined as:

$$err = y - \hat{y}lin \qquad (2)$$

The expected prediction error was therefore modelled as a smooth function of kidney coordinates:

$$\hat{e} = f(eGFR, \text{log-transformed UACR}), \qquad (3)$$

where *f* was a spline-Ridge regressor fitted in the kidney-diverse cohort using cubic B-spline bases (four knots per axis, degree 3, uniform knots, constant extrapolation, Ridge α = 1·0, no cross-terms), median imputation of missing kidney inputs, and a minimum of 80 eligible observations per surface.

The resulting error surface was used to correct UPBioAge:

$$\hat{y}k\text{-}UPBioAge = \hat{y}UPBioAge + \hat{e}\_applied, (4)$$

where ê_applied is the correction after the gate defined in equation (5), yielding the kidney-conditioned k-UPBioAge clock. Positive correction values increase the estimated age of samples for which kidney-domain structure would otherwise bias UPBioAge downward.

**Dual-arm calibration procedure.** The UPBioAge derivation cohort was restricted to preserved kidney function (Table 1), providing insufficient kidney-function variability to estimate the relationship between prediction error and kidney function. FAC was therefore trained in the independent kidney-diverse cohort.

To enable application both when kidney measurements were available and when they were absent, calibration proceeds in three steps. First, the two kidney coordinates are resolved independently of each other: eGFR and UACR are each taken from the measurement where one exists and from the peptide-derived estimate otherwise, so a participant may carry a measured eGFR alongside an estimated UACR. Second, two calibration surfaces were developed. The measured-coordinate surface was fitted in the 7,448 kidney-diverse participants who have a measured eGFR and a recorded age (of the 7,540 with a measured eGFR), and was supplied with the resolved albuminuria coordinate rather than a measured one (the measured ratio in 4,620 of them and the peptide-derived estimate in the other 2,828; a further 254 participants had a measured UACR but no measured eGFR), which is exactly the mixture of measured and estimated coordinates it receives when it is applied. The second surface was fitted on peptide-derived estimates of both coordinates in the 7,704 participants with a recorded age. Each surface is therefore trained on the same kind of input it subsequently receives, including for participants who have a measured eGFR but no measured albuminuria. A participant is scored by the surface matching whether a measured eGFR was available, so in the clinical follow-up cohort, where eGFR was measured in all 7,469 participants, every participant was scored by the measured-value surface. Third, the resulting correction is set to zero inside the KDIGO low-risk zone and retained outside it: it is kept where the resolved eGFR is below 60 mL/min/1·73 m², or the resolved UACR exceeds 30 mg/g, or no resolved eGFR can be formed at all, that is, neither a measured value nor a peptide-based estimate is available, and set to zero elsewhere.

$\hat{e}$_applied = $\hat{e}$ *if eGFR < 60 mL/min/1·73 m², or UACR > 30 mg/g, or no resolved eGFR exists;*
$\hat{e}$_applied = 0 *otherwise (5)*

The third condition is a safeguard and applies to no participant in this study, because every participant has at least a peptide-based estimate. The albuminuria arm of the gate is written as a strict inequality, whereas KDIGO category A2 begins at 30 mg/g; no participant in the clinical follow-up cohort has a resolved UACR of exactly 30 mg/g, so the two conventions classify every observation identically. In the clinical follow-up cohort this leaves the prediction unchanged for 3,649 of 7,469 participants and applies a correction to the remaining 3,820. Because the gate is a threshold, the applied correction is discontinuous at its boundaries: for normoalbuminuric participants it steps by about 3 years at an eGFR of 60 mL/min/1·73 m², and at a UACR of 30 mg/g by about 2 years.

UPBioAgeAcc represents total urinary peptidomic age acceleration, encompassing both biological ageing and kidney-associated variation in the urinary peptidome. k-UPBioAgeAcc represents the corresponding age acceleration after conditioning on eGFR and UACR.

The kidney-attributable share of variance in an age acceleration measure was quantified as the $R^2$ of an ordinary least-squares regression of that measure on the two kidney coordinates the calibration uses (eGFR and ln(1 + UACR), each taken as the measured value where one exists and the peptide-based estimate otherwise), fitted in the clinical follow-up cohort (n = 7,469) before and after conditioning, with the signs of the regression coefficients giving the direction of the association. FAC does not remove kidney-associated information. Rather, it re-orients this component of the age signal: the proportion of variance attributable to kidney function increased from 1·5% to 11·9%, and the albuminuria coefficient changed sign from negative to positive while the eGFR coefficient remained positive. k-UPBioAgeAcc therefore remains kidney-responsive and should be interpreted as kidney-conditioned rather than causally independent of kidney function.

**Control analyses.** To determine whether FAC reintroduced chronological-age information into the calibrated prediction, the individual correction ($\hat{e} = \hat{y}$k-UPBioAge − $\hat{y}$UPBioAge) was regressed on chronological age in every cohort, rather than only in the derivation set, where age-orthogonality of age acceleration holds by construction.

Chronological age explained 7·6% of correction variance in the derivation OOF set (slope +0·010 correction-years per chronological year) and 10·6% in the internal holdout (slope +0·013). Out of sample it explained 28·1% in the Glasgow samples (slope +0·114; n = 675), 12·1% in FLEMENGHO (slope +0·035; n = 778), and 6·2% in the clinical follow-up cohort (slope +0·069); the slopes remain small, at most about one-tenth of a year of correction per year of age.

The FAC correction therefore contains a small but non-zero age component where kidney function itself varies with age, as expected for a correction derived from eGFR and albuminuria. This further supports interpreting k-UPBioAgeAcc as kidney-conditioned rather than kidney-independent and is why all reported outcome models were additionally adjusted for chronological age.

**Supplementary Methods S5: Mortality analysis**

**Model checks and sensitivity analyses.** Proportional hazards for k-UPBioAgeAcc were assessed by scaled Schoenfeld residuals of that term under rank, log, and Kaplan–Meier time transformations (all-cause mortality p=0·32, 0·20, and 0·25); these are tests of the score, not global tests of every covariate; where the assumption held, Cox HRs are interpreted as constant associations over follow-up. The albuminuria axis was adjusted in three ways, all reported in Supplementary Table S10: with the measured UACR in the 2,482 participants (33%) who have one, which is the primary analysis; with the value the calibration itself applies in every participant, which is the measurement where one exists and a peptide-based estimate otherwise; and with the peptide-based estimate in the 4,987 participants who have no measurement. As a check on reverse causation, follow-up was partitioned at the median landmark (3·95 yr) and the association re-estimated separately before and after that landmark; the resulting HRs are reported in the main text. The same model was refitted three times after excluding participants who died within 0·5, 1, and 2 years of urine sampling, so that the deaths closest to sampling, those most likely to reflect illness already present at the time the urine was taken, could not drive the association; these HRs are also given in the main text. Cumulative-incidence curves used tertile boundaries with a number-at-risk table and the k-group log-rank statistic. Sex- and age-group-stratified survival analyses (Supplementary Table S9) tested consistency across demographic strata. Harrell's C-index and ΔC, the continuous net reclassification improvement (NRI) and the integrated discrimination improvement (IDI) (all reported in Supplementary Table S12) were all computed OOF, using stratified cross-validation with five folds repeated five times, so that every participant's predicted risk came from a model that had not been fitted on them, with 95% confidence intervals from 500 bootstrap resamples. NRI and IDI were evaluated at a 5-year horizon. Every covariate used in the outcome models, that is chronological age, sex, body-mass index (BMI), mean arterial pressure, measured eGFR, and the four comorbidity indicators, was available for all 7,469 participants, so the fully adjusted models are complete-case analyses of the whole cohort with no covariate-driven exclusion. Of those 7,469 participants, 625 died during follow-up and 6,844 were censored, the longest observed follow-up being 20·2 years. Follow-up was conducted by the contributing studies, which supplied an observed time and an event indicator, so participants lost to follow-up cannot be distinguished from those alive at the close of each study and no comparison between the two is possible.

**Study-level and quintile analyses.** The incident endpoints were ascertained in a subset of studies with too few events per study for stable study-specific estimates, and were therefore analysed in the pooled cohort only. As a sensitivity analysis, the fully adjusted model for all-cause mortality was also refitted stratified by contributing study, allowing each study its own baseline hazard. Quintile analyses of both age acceleration measures were adjusted for age and sex, with the lowest quintile as the reference, and the median measured eGFR and the proportion with a measured albuminuria of at least 30 mg/g are reported for each group. Each quintile was compared with the lowest quintile in a separate age- and sex-adjusted Cox model restricted to those two groups.

**Incremental prognostic value.** To assess whether k-UPBioAgeAcc provided additional prognostic information for all-cause mortality beyond the variables included in the fully adjusted model, we compared the fully adjusted Cox model with and without k-UPBioAgeAcc. Improvement in model fit and discrimination was assessed using the likelihood-ratio test, difference in Akaike information criterion, and change in Harrell's C-index. Continuous net reclassification improvement and integrated discrimination improvement at 5 years were additionally evaluated. These measures answer different clinical questions: Harrell's C-index asks only whether patients are ranked in the right order, whereas the reclassification measures ask whether, and by how much, an individual's estimated risk moves in the right direction.[12,13] All three measures were estimated by cross-validation, with bootstrap confidence intervals.

**Supplementary Methods S6: Incident events and disease-burden age acceleration**
**Incident-event analysis.** Incident cardiovascular and renal events were taken from the adjudicated endpoint definitions of Jaimes Campos et al.[14] The composite cardiovascular endpoint and its CAD and HF components are defined as fatal or non-fatal, so a death is counted within the endpoint rather than alongside it: among the 5,560 participants followed for cardiovascular events, all 501 deaths coincided with a recorded CAD or HF event, and none of the 4,726 event-free participants was recorded as having died, so deaths make up 501 of the 834 composite events (179 of the 363 CAD and 322 of the 471 HF events). The 5,560 participants come from the 12 contributing studies that followed cardiovascular events; uPROPHET, PersTIgAN, Predictions (HIB), STOP-IgAN, and TransBioBC contributed no cardiovascular follow-up. CKD progression, defined as a decline in eGFR of at least 40%, is not death-defined: none of the 113 participants with an event died. It was followed in 3,617 participants from six studies (DIRECT, EPOGH, EU-PRIORITY, Heart failure, Predictions [Prague], and Sun-MACRO). Each participant contributes a first cardiovascular event only. The CAD and HF groups are therefore mutually exclusive, no participant appears in both, and the composite count is exactly their sum (363 + 471 = 834). The two component endpoints are analysed as cause-specific first-event outcomes on a single common risk set: all 5,560 participants followed for cardiovascular events enter at the time of urine sampling, and a participant whose first event was of the other type is censored at that event rather than removed from the analysis. Entry is therefore never conditioned on which event a participant went on to have, and the CAD, HF, and composite analyses all share the denominator of 5,560. The component estimates should still be read as analyses of the first such event rather than of any occurrence of each condition.

**Competing risks for CKD progression.** CKD progression is defined by a decline in eGFR rather than by death, so deaths can remove participants from its risk set: 66 participants died without a CKD event, against 113 events, although only 10 of these deaths occurred within CKD follow-up and the other 56 after the last kidney-function measurement. This endpoint was therefore also examined under a Fine–Gray subdistribution model adjusted for the same covariates, counting all 66 deaths as competing events, in which the estimate was similar in magnitude (subdistribution HR 1·37 per SD, 95% CI 1·04–1·81, p=0·027, against a cause-specific HR of 1·36, 95% CI 1·08–1·72, in the identically standardised model). Counting only the 10 deaths within follow-up as competing events gave a similar estimate (subdistribution HR 1·36, 95% CI 1·02–1·81, p=0·034). The estimate reported for CKD progression in Supplementary Table S8 remains the cause-specific one. Both subdistribution models were fitted with the finegray function of the R survival package (version 3.8.6) and a weighted Cox model with participant-clustered variance. All 113 progression events occurred in participants with diabetes, so the diabetes coefficient is not estimable (it diverges, which the software reports as a convergence warning); refitting both models in the 3,095 participants with diabetes, without that term, gave the same estimates (1·37, 95% CI 1·03–1·81, and 1·36, 1·02–1·81). That table reports 1·39 (1·08–1·78) for the same endpoint and covariates: the two differ only in how the age acceleration measure is standardised, the log HRs being a constant multiple of one another, and not in the participants, events, or model.

**HRs and cumulative incidence.** The four secondary endpoints were not adjusted for multiplicity in the models; Holm correction across them is reported in the main text. Every HR reported in the tables and figures comes from a cause-specific Cox model, which is itself a standard way of analysing competing-risk data and estimates the rate of the event among those still at risk. Three distinct procedures are therefore used and should not be conflated: HRs from cause-specific Cox regression; cumulative incidence for display, estimated as one minus the Kaplan–Meier estimate for all-cause mortality and with the Aalen–Johansen estimator for the incident endpoints, where death is treated as a competing event; and comparison of tertiles by the log-rank test, which compares the cause-specific hazards between tertiles rather than the displayed cumulative-incidence curves themselves.

**Disease-burden age acceleration.** Disease-burden age acceleration was quantified by ordinary least-squares models of the form age acceleration ~ disease pattern + chronological age + sex + BMI + mean arterial pressure across all sixteen combinations of diabetes, cardiovascular disease, CKD, and hypertension, with the comorbidity-free group as reference; reported values are disease-pattern coefficients (years, 95% CI) with two-sided P from the coefficient t-test. The in-cohort age and sex terms absorb residual out-of-sample age dependence in age acceleration, while BMI and mean arterial pressure are adjusted for as clinical confounders; none of these terms modifies the k-UPBioAgeAcc variable itself. The number of participants is reported for every combination; mortality follow-up was not used.

**Accuracy by comorbidity stratum.** We deployed the calibrated clock across the clinical follow-up cohort stratified into six mutually exclusive comorbidity strata defined on the recorded diagnoses: a comorbidity-free reference stratum (no recorded diabetes, kidney disease, hypertension, or cardiovascular disease [CVD]), diabetes-only, kidney-disease-only, hypertension-only, CVD-only, and multiple-comorbidity; the comorbidity-free reference stratum comprises 690 participants. Because the unconditioned and the kidney-conditioned age acceleration are measured in the same participants, the effect of conditioning within each stratum was quantified as a paired effect size, Cohen's dz = mean(k-UPBioAgeAcc − UPBioAgeAcc) / SD(difference). Prediction accuracy within each stratum is reported against the same metrics used for the external cohorts. Stratum sizes, accuracy, and the resulting paired effect sizes are reported in Supplementary Table S7.

**Supplementary Methods S7: Sensitivity and robustness analyses**

**Why the unconditioned clock requires kidney conditioning.** Ranking participants by unconditioned age acceleration produced a biologically implausible ordering: median measured eGFR rose steadily across quintiles of apparent biological age, so the quintile identified as most aged had the best-preserved filtration, and the proportion with albuminuria of at least 30 mg/g fell across the same quintiles. Conditioning reversed both orderings and restored a monotone mortality gradient (Supplementary Figure S6). Within strata of measured eGFR, adjusted for age and sex, the mortality HR per within-stratum SD of the unconditioned measure fell from 1·36 and 1·37 in the two highest eGFR categories to 1·22, 1·10, and 0·97, so that in KDIGO stages G4–G5 it showed no association, whereas k-UPBioAgeAcc generally retained stronger associations across the same strata, although no association was evident in G4. Selecting on eGFR alone does not identify the participants in whom the correction is switched off, because those with albuminuria above 30 mg/g still receive one. Because the correction is confined by construction to participants outside the KDIGO low-risk zone, Figure 2B of the main text reports k-UPBioAgeAcc itself across the KDIGO grid, with the pooled low-risk zone as reference.

**Quintiles of the unconditioned measure.** Dose-response is lost among the participants whose kidneys look healthy: among those with an eGFR of at least 60 mL/min/1·73 m², quintiles 2 to 4 of the unconditioned measure carried HRs of 1·34 (95% CI 1·00–1·80), 1·30 (0·94–1·82) and 1·31 (0·89–1·93) relative to the lowest quintile, flat through the middle of the distribution with every interval touching 1, before rising to 3·01 (1·87–4·84) in the top quintile. After FAC the gradient is restored in the same participants, monotone from the second quintile upwards (1·39, 1·47, 1·98, and 4·10). Across the whole clinical follow-up cohort the most aged quintile carried an HR of 3·66 (2·81–4·75) after conditioning against 2·32 (1·75–3·08) before it, each relative to the least aged quintile and adjusted for age and sex.

**Conditioning is not interchangeable with adjustment.** Among the 1,905 participants with a measured eGFR below 60 mL/min/1·73 m², a Cox model already containing a natural cubic spline of measured eGFR showed no association for the unconditioned measure (HR 1·05 per SD, 95% CI 0·93–

1·20; p=0·42), whereas k-UPBioAgeAcc remained associated (1·22, 1·06–1·40; p=0·0059). Where filtration is impaired, the distortion therefore appears to lie in the score itself rather than in the covariate set of the outcome model. Conditioning does not remove kidney biology from the marker: it re-orients kidney-associated variation so that variation which previously ran counter to mortality risk now runs with it (variance decomposition in Supplementary Methods S4). Albuminuria contributed most strongly to the conditioning, with eGFR adding information, consistent with their different effects on urinary peptide composition. In the same participants the fully adjusted HR for k-UPBioAgeAcc exceeded that for the unconditioned measure by a factor of 1·21 (95% CI 1·16–1·27; p<0·001, paired bootstrap), and in no resample did the unconditioned measure perform better.

**Recovered ageing signal or imported kidney information?** Because FAC adds a function of eGFR and albuminuria to the clock, a stronger outcome association after conditioning could reflect recovery of a distorted ageing signal, incorporation of kidney-related prognostic information, or both. To separate these, the two acceleration measures were compared on identical participants against a model that already contains kidney function flexibly. In the 2,482 participants who have both kidney markers measured (200 deaths), a Cox model containing age, sex, BMI, mean arterial pressure, diabetes, cardiovascular disease, hypertension, and natural cubic splines of measured eGFR and of log measured UACR reached a C-index of 0·885. Adding UPBioAgeAcc gave an HR of 1·29 per SD (95% CI 1·09–1·54; likelihood-ratio p=0·003, ΔAIC −6·6, ΔC +0·0003) and adding k-UPBioAgeAcc an HR of 1·32 per SD (1·11–1·57; likelihood-ratio p=0·001, ΔAIC −8·2, ΔC +0·0006). Both measures therefore retain an association with mortality beyond flexibly modelled kidney function, and the difference between them narrows considerably once kidney function is modelled flexibly rather than as a single linear term, which is consistent with part of the whole-cohort advantage of conditioning being kidney-related prognostic information rather than recovered ageing signal alone. The correction term itself carries little independent information: being a smooth function of the two resolved kidney coordinates, it is reproduced with $R^2 = 0·95$ by additive splines of those coordinates, so it cannot add to a model that already contains them. This comparison is, however, confined to the participants in whom albuminuria was measured, and that subgroup does not span the kidney spectrum: 95·5% of them have an eGFR of at least 60 mL/min/1·73 m², and measured albuminuria is available for only 9 of the 473 participants in KDIGO stages G4–G5. Most of them therefore lie inside the low-risk zone, where no correction is applied at all, so this analysis cannot establish what conditioning does where filtration is impaired. Where filtration is impaired, flexible modelling of kidney function in the outcome model does not substitute for conditioning (see Conditioning is not interchangeable with adjustment, above). In advanced CKD, the two explanations cannot be fully separated with these data, because measured albuminuria is almost entirely absent there.

**Attenuation by kidney measures.** Adding eGFR to a model adjusted for age, sex, BMI, diabetes, cardiovascular disease, and hypertension (HR 1·53) attenuated the mortality association by 10·2% (95% CI 0·3–19·3). Adding albuminuria as the calibration uses it to that model plus eGFR attenuated it by 37·8% (95% CI 25·3–51·7; HR 1·27, 95% CI 1·15–1·40); attenuation was 50·0% among the 2,482 participants with a measured UACR, in a model omitting cardiovascular disease, which is near-separating in that subset (HR 1·34; 95% CI 1·12–1·61; 31·3% with it retained), and 34·9% among the 4,987 without one. The mortality signal was therefore not fully explained by albuminuria in any of the three analyses (Supplementary Table S10).

**Attenuation method.** The eGFR attenuation analysis used a difference-method Cox decomposition (n = 7,469, measured eGFR for all), with attenuated proportion = (k-UPBioAgeAcc_alone − k-UPBioAgeAcc_adjusted-for-eGFR)/k-UPBioAgeAcc_alone and 400-bootstrap CIs (Supplementary Table S10). The attenuated proportion is interpreted as a statistical partitioning of association, not as a causal natural direct or indirect effect.

**The zero-correction subgroup.** Inside the KDIGO low-risk zone no correction is applied, so the two predictions are numerically identical. The two acceleration measures are not quite identical even so, because each is the residual of its own prediction on chronological age using its own fixed coefficients; within that subgroup they differ by a mean of −0·18 years (range −0·59 to +0·17), and the HRs they give agree to three decimal places. A correction derived from kidney measurements could in principle generate the association it appears to reveal, but with no kidney-derived term entering the marker in this subgroup (3,649 of 7,469 participants), the association with mortality was still present under full adjustment (HR 1·33, 95% CI 1·02–1·74 per SD, 118 deaths, p=0·035) and did not differ from that among participants who did receive a correction (P for interaction 0·20; Supplementary Table S14), so within that subgroup the association cannot be attributed to the correction.

Restricting the zero-correction subgroup to the 2,117 participants who have both kidney markers actually measured, rather than one of them estimated, leaves the estimate in the same direction with a wider interval and 73 deaths (HR 1·25, 95% CI 0·89–1·74, p=0·20).

There was also no statistically detectable interaction according to whether albuminuria had been measured directly or estimated from the peptide data (P for interaction 0·39; Supplementary Table S14), although the subgroup estimates themselves differed in size.

**Strata without an association.** In the smallest and most severely affected strata the mortality association was not evident: at an eGFR of 15–29 mL/min/1·73 m² (KDIGO G4; n = 433, 84 events; HR 0·93, 95% CI 0·71–1·22) and at macroalbuminuria of at least 300 mg/g on a measured UACR (n = 92, 34 events; HR 0·86, 95% CI 0·56–1·31). These HRs are per SD of k-UPBioAgeAcc computed within each stratum, from the fully adjusted model without its eGFR term. These two strata differ in what can be read from them. The G4 stratum could have detected any HR at or above 1·47 per within-stratum SD with 80% power, so its null estimate is informative; the macroalbuminuria stratum was limited to HRs at or above 1·85. The stratum with an eGFR below 15 mL/min/1·73 m² informs the question neither way: with 40 participants and 14 deaths its estimate is 1·62 (95% CI 0·60–4·36), consistent with the whole-cohort association but detectable in that stratum only at 4·10 or above. The macroalbuminuria subgroup is defined on measured albuminuria only, which is why it is much smaller than the 1,002 participants placed in KDIGO A3 in Figure 2B, where the category uses the resolved coordinate and is therefore an estimate in most of them. In participants this far into kidney failure, mortality risk is plausibly dominated by filtration failure itself, leaving little residual variance for an ageing signal to track independently. For G4 the present data do distinguish the two explanations; for the two smaller strata they do not.

**Interaction tests and power.** Disease-specific interaction tests added multiplicative k-UPBioAgeAcc × {diabetes, cardiovascular disease (CVD), hypertension, kidney disease} terms individually to a model adjusted for age, sex, and the four comorbidity indicators, with BH correction across the four interaction tests, accompanied by a matched-power check for the CVD-positive and CVD-negative strata (two-sided $\alpha = 0{\cdot}05$, target HR 1·25), to distinguish genuine within-disease enrichment from differential power. Using the standard error observed in each stratum, the minimum HR detectable with 80% power is 1·19 in the cardiovascular-disease-positive stratum (374 deaths) and 1·25 in the cardiovascular-disease-negative stratum (251 deaths); both strata were therefore powered to detect the target HR of 1·25, so the similar estimates in the two strata (1·51 and 1·35) are not a power artefact.

**Between-study consistency.** A per-study DerSimonian–Laird random-effects meta-analysis pooled the 12 contributing studies each contributing ≥10 deaths (Supplementary Table S1, forest plots in Supplementary Figure S5). Where only one participant of a study differed on a yes/no covariate, that covariate was omitted from the study's model (cardiovascular disease in HOMAGE-Fibrosis, for both measures). Consistency was checked further by refitting the fully adjusted model once for each of the 17 contributing studies of Supplementary Table S1, each time excluding one study: the HR per SD ranged from 1·39 to 1·52 with a mean of 1·47, against 1·47 in the whole cohort, so no single study drives the association. CKD-stage stratification was fitted within KDIGO stages with ≥10 deaths (Supplementary Table S9).

**Judging a conditioned clock.** Because conditioning deliberately re-orients the variation that tracks chronological age through kidney function rather than leaving it in place, a conditioned clock is not expected to agree more closely with chronological age than an unconditioned one. Agreement with chronological age and association with outcome test different properties of a clock, and neither alone identifies which measure better indexes ageing. The appropriate test is whether the departure from chronological age carries prognostic information.

**Supplementary Methods S8: Enrichment, statistics, reproducibility, and supporting detail**

**Enrichment.** Over-representation analysis was run with g:Profiler (gprofiler-official, organism hsapiens) on the gene symbols of the parent proteins of the retained peptides, restricted to Gene Ontology (GO) Biological Process, Reactome, and the Kyoto Encyclopedia of Genes and Genomes (KEGG). Of the 685 parent proteins of the 2,631 retained peptides, 667 carried a gene symbol and 657 mapped to the g:Profiler namespace and formed the query. The domain scope was a custom background of the CE–MS-detectable urinary proteome derived from the peptide library (effective domain 2,448 proteins), so enrichment is assessed against what this assay can observe rather than the whole genome; significance was a BH false discovery rate of 0·05. The query proteins are the parent proteins of model peptides rather than an independently selected gene set, so the analysis is descriptive and no causal or mechanistic claim follows from it. The background comprised the parent proteins of all library peptides, including proteins with no peptide passing the detection threshold, and the analysis does not account for the differing numbers of eligible peptides per parent protein, which give proteins represented by many peptides, the collagens in particular, more opportunities to enter the query. The results reported are from the query of 8 October 2026 (the 12 October 2026 run returned an identical result); the query, background and complete result table are archived with the code, because the annotation databases behind the service are updated over time.

**Composition of the retained peptides.** Collagen alpha-1(I) contributed the largest number of fragments and collagen alpha-1(III) the largest net age-positive contribution. Ageing is therefore marked by a change in which collagen fragments appear in urine rather than by more or less collagen overall. Several peptide-age relationships were also clearly non-linear, in line with the non-monotonic trajectories reported in plasma and multi-omic proteomic studies.[6] Fibrinogen alpha- and beta-chain fragments offer a link to coagulation, acute-phase responses, and inflammaging.[15,16] Twenty-two polymeric immunoglobulin receptor fragments together formed the strongest net non-collagen contribution and suggest an epithelial and mucosal transport component.

**Renal components of the clock.** Renal processes are also represented in the model. Eight uromodulin peptides were retained; uromodulin is produced in the thick ascending limb and reflects tubular integrity and functional nephron mass.[17,18] Clusterin fragments point to renal epithelial stress and senescence, and urinary clusterin has been associated with CKD progression after adjustment for eGFR, albuminuria, age, and sex.[19] Peptides of this kind would have been removed by an exclusion-based approach. Conditioning is directed at variation attributable to filtration and barrier integrity, not at the ageing kidney itself.

**Generalisation to the Glasgow cohort.** Age prediction also held in the independent Glasgow cohort (n = 675; MAE 5·44 years; r = 0·74), whose samples were analysed in a different laboratory on an analogous CE–MS platform. This cohort also differed geographically: 604 of its 675 samples (89%) came from South African study populations, although ethnicity was not recorded in them either. Transferability between laboratories is therefore supported, although transfer to other analytical platforms has not been examined.

**Discrimination in context.** The pattern of a mortality association that persists after clinical adjustment alongside a modest discrimination gain is consistent with epigenetic and plasma proteomic ageing clocks.[7,20,21] Epigenetic age acceleration has repeatedly been associated with mortality, including in a meta-analysis of 13 cohorts in which each additional year of extrinsic epigenetic age acceleration carried about a 4% higher risk of death,[22] while GrimAge is among the strongest methylation-based predictors of mortality and lifespan, carrying about a 10% higher risk of death for each additional year of age acceleration.[7] Plasma proteomic ageing markers have similarly been associated with mortality, multimorbidity, and a broad range of incident age-related diseases.[21,23] These should not be regarded as competing measures, because they capture partly different components of biological ageing. Plasma proteomic clocks rest mainly on circulating protein abundance, whereas urinary peptidomics predominantly measures fragments generated by proteolytic processing and subsequently filtered or released into urine. The urinary peptidome therefore extends the set of molecular compartments from which biological ageing can be read. Urinary miRNA-based predictors offer a further non-invasive route,[24] indicating that different molecular layers within urine capture distinct aspects of ageing.

A comparable example in another ageing marker is a cohort of African American adults followed for incident cardiovascular disease, in which adding epigenetic age acceleration to age, sex, and an established cardiovascular risk score moved the C-index from 0·687 to 0·698, and from 0·670 to 0·685 against a second risk score; neither increase reached statistical significance, yet the reclassification of individual risk still improved.[25]

In the present, larger cohort, the corresponding gain did reach statistical significance; the outcome, the baseline model, and the population all differ, so the magnitudes should not be read as equivalent.

**Precedent for adjusting an ageing marker.** Adjusting an ageing marker for a known source of distortion has precedent. Intrinsic epigenetic age acceleration is derived after accounting for chronological age and blood cell composition,[22,26] and neuroimaging brain age estimates are corrected for systematic demographic and acquisition-related effects.[9,27] The urinary case differs from these examples in an important respect. In those settings the distorting factor is a property of the population or of the instrument, whereas kidney function determines the molecular composition of the sample itself.

**Excluding kidney-associated peptides instead.** A previous urinary peptidomic cancer classifier handled kidney-related peptides by excluding those overlapping the predefined CKD273 classifier.[28] We instead retained the complete ageing peptidome and conditioned the resulting age acceleration on kidney function, because kidney-associated peptides can themselves carry information about biological ageing. To our knowledge this is the first urinary ageing clock in which kidney function is explicitly accounted for.

**Biofluid gating.** Urine is the first case of this problem rather than the general one. A biofluid generated by an organ can carry that organ's functional state into the measurements it yields: saliva through glandular flow, stool through transit time and microbial mass, cerebrospinal fluid through barrier integrity, plasma for any analyte cleared by the kidney. Wherever that holds, the function of the producing organ may need to be treated as a calibration axis of the assay and conditioned on before the measurement is read as systemic biology, rather than added as one more covariate, because a covariate cannot restore information the organ has already removed. We call this principle biofluid gating, and propose it as a testable design principle rather than a settled requirement: this study tests it for urine and the kidney only.

**Statistics and reproducibility.** Prediction accuracy was summarised as MAE, RMSE, Pearson r, and $R^2$. Calibration was assessed as calibration-in-the-large, calibration slope, and age-binned residuals, following Van Calster et al,[29] because a model that ranks people well can still be systematically off in absolute terms when it is applied to a new cohort. Groups were compared with two-sided Welch t-tests, Mann–Whitney U tests, and Cohen's d. No statistical method was used to predetermine sample size; all eligible archived samples were used. The resulting outcome cohort of 7,469 participants with 625 deaths provides approximately 78 events per adjustment covariate in the fully adjusted model (69 per estimated coefficient, including k-UPBioAgeAcc), and the minimum detectable effect size is reported alongside each analysis for which power is a material consideration. Random seeds were fixed and are recorded in the analysis scripts: 42 for the primary cross-validation, 104 for the attenuation bootstraps, 20260813 for the kidney-surrogate search, and 20260819 for the paired hazard-ratio bootstraps. Cox models for Supplementary Tables S8 and S11 were fitted with statsmodels (PHReg, Efron ties); lifelines was used for the proportional-hazards tests, the attenuation bootstraps, the stratified and sensitivity models, and the cross-validated discrimination measures; Fine–Gray models were fitted in R (survival 3.8.6).

**Code and data availability.** Code to reproduce the model fitting, calibration, and statistical analyses, together with the complete fitted model, is deposited on Zenodo (doi:10.5281/zenodo.22914811); editors and reviewers are given access during peer review, and the deposit will be made public on publication. The deposit contains the complete pipeline, the fitted model, and a requirements file specifying the software environment given in Supplementary Methods S3, with a single-command reproducibility entry point. The deposit excludes proprietary database credentials and participant-level data, which remain available under data-use agreements (see Data sharing in the main text).

**Implementation.** Each CE–MS run passes the laboratory's routine quality control before its data are used, including calibration of mass, migration time and signal intensity against internal standard peptides and regular analysis of a standard urine sample;[3] all samples in this study passed these procedures, and a sample that fails them should not be scored. Peptides below the detection threshold are recorded as zero and handled by the model as the lowest abundance (Supplementary Methods S3), so no imputation is needed. CE–MS acquisition and processing require a specialist laboratory; the model itself is fully scripted, with no user-adjustable parameters, and clinical eGFR and albuminuria are used where available and estimated from the peptide data otherwise.

**Supplementary Methods S9: In-silico intervention analysis**

This analysis reports a simulation and not observed treatment responses. The peptide-level effects of each intervention were taken from previously published urinary peptidomic intervention studies, in which the change in individual peptides under a given treatment was quantified; they were not generated here. The procedure for carrying measured peptide changes through a peptide-based score is the one established by Jaimes-Campos and colleagues,[14] and the per-peptide factors for all six interventions were taken as one set from the compiled intervention table of the in-silico multifactorial-intervention study of Latosinska and colleagues,[30] which assembled them from those primary studies. The six interventions are: sodium–glucose cotransporter-2 (SGLT2) inhibition, exercise, angiotensin-receptor blockade, spironolactone, glucagon-like peptide-1 (GLP-1) receptor agonism and olive oil. Each intervention is represented as a multiplicative factor per peptide, covering 920 peptides of the age panel; every other peptide in the model is carried through unchanged, which is equivalent to a factor of one. These values are model projections. They are not evidence that any of these treatments alters biological ageing, and they were not used in any other analysis in this paper. Whether k-UPBioAge changes as projected under actual treatment will be tested against measured treatment responses in a dedicated follow-up study.

**Applying the factors, and participant selection.** For every participant in the clinical follow-up cohort the factors for one intervention were applied to that participant's raw peptide abundances, and the complete clock was then re-run on the modified profile: stage 1, stage 2, linear recalibration, the spline correction, and FAC. Everything the peptides determine is therefore recomputed, including the peptide-based eGFR and albuminuria estimates and the calibration correction that follows from them. Measured kidney values are not altered by the simulation, so in the clinical follow-up cohort, where eGFR is measured in every participant and albuminuria in 2,482, those measured coordinates stay at their baseline values and only the peptide-derived coordinates move. The predicted change is the difference in k-UPBioAge between the modified and the baseline profile, in years. The analysis covers all 7,469 participants and all six interventions and used the kidney surrogate models fitted before two erroneous eGFR records were corrected; the corresponding figure displays ten of those participants, labelled P1 to P10, in ascending order of baseline k-UPBioAge. They were selected by a fixed rule rather than for their predicted response: the participant nearest each of the 5th, 15th, 25th, 35th, 45th, 55th, 65th, 75th, 85th, and 95th percentile of baseline k-UPBioAge across all 7,469.

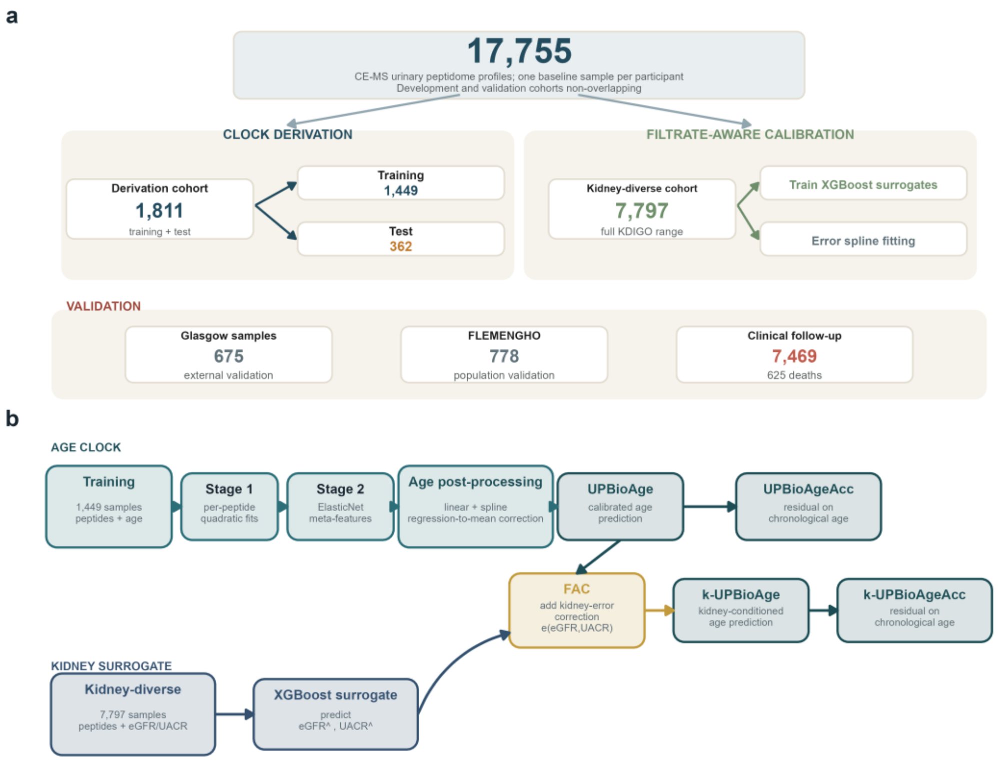


**Supplementary Figure S1 | Study cohorts and analytical workflow.**

a, Overview of study cohorts to derive, kidney-condition (by FAC) and validate the urinary peptidomic ageing clock. The analysis comprised 17,755 CE–MS urinary peptidome profiles (the named cohorts total 18,530 memberships because 775 FLEMENGHO participants also belong to the clinical follow-up cohort and are counted once in each. FLEMENGHO has no row of its own in Supplementary Table S1: those 775 appear there under the studies through which they were sampled, 714 under EPOGH and 61 under Heart failure). One baseline sample per participant was used. The derivation cohort (n = 1,811) was divided into training (n = 1,449) and internal holdout (n = 362) sets. A kidney-diverse cohort spanning the KDIGO range (n = 7,797) was used to train the eGFR and UACR peptide surrogates and fit the FAC error surface. Independent validation included the Glasgow cohort (n = 675), the FLEMENGHO cohort (n = 778) and the clinical follow-up cohort (n = 7,469). b, Analytical workflow. The age clock was trained in a two-stage approach. First, per-peptide quadratic age predictions were calculated and used as input to the stage 2 elastic-net model. Post-processing involved linear recalibration followed by spline regression-to-the-mean correction, yielding UPBioAge. In the kidney-diverse cohort, the peptide-based eGFR and UACR surrogates were used by FAC to estimate and re-orient kidney-associated error, and FAC was applied to UPBioAge to give the kidney-conditioned prediction k-UPBioAge. Each acceleration measure is the residual of its own prediction on chronological age: UPBioAgeAcc from UPBioAge, and k-UPBioAgeAcc from k-UPBioAge. CE–MS, capillary electrophoresis–mass spectrometry; eGFR, estimated glomerular filtration rate; FAC, Filtrate-Aware Calibration; FLEMENGHO, Flemish Study on Environment, Genes and Health Outcomes; KDIGO, Kidney Disease: Improving Global Outcomes; UACR, urinary albumin-to-creatinine ratio; k-UPBioAge, kidney-conditioned urinary peptidomic biological age.

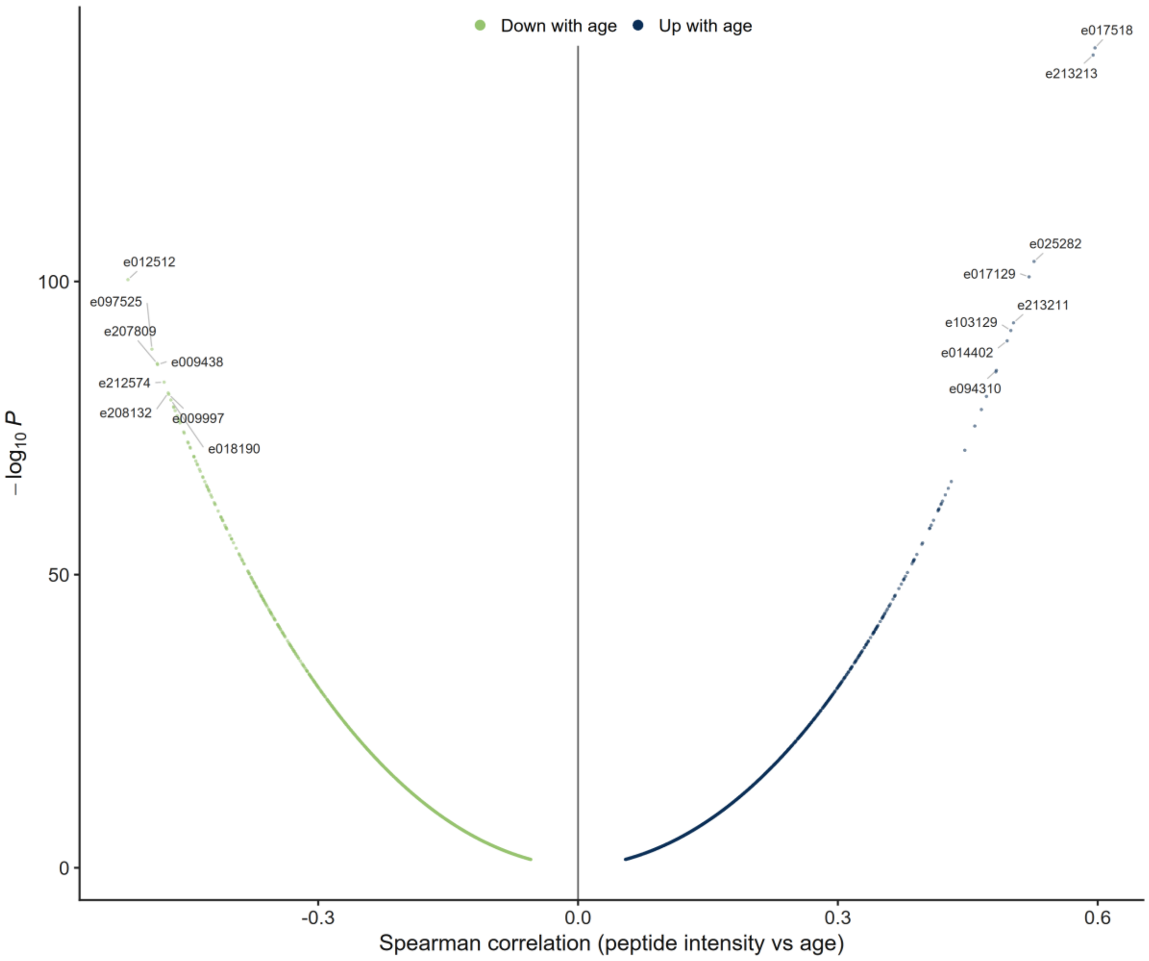


**Supplementary Figure S2 | Per-peptide age-correlation volcano.**

Spearman rank correlation with chronological age in the training set for the 3,296 peptides significantly associated with age after Benjamini–Hochberg correction (q < 0·05), the screen that defined the stage-2 input; x = correlation coefficient, y = −log10 P; colour shows the direction of the correlation.

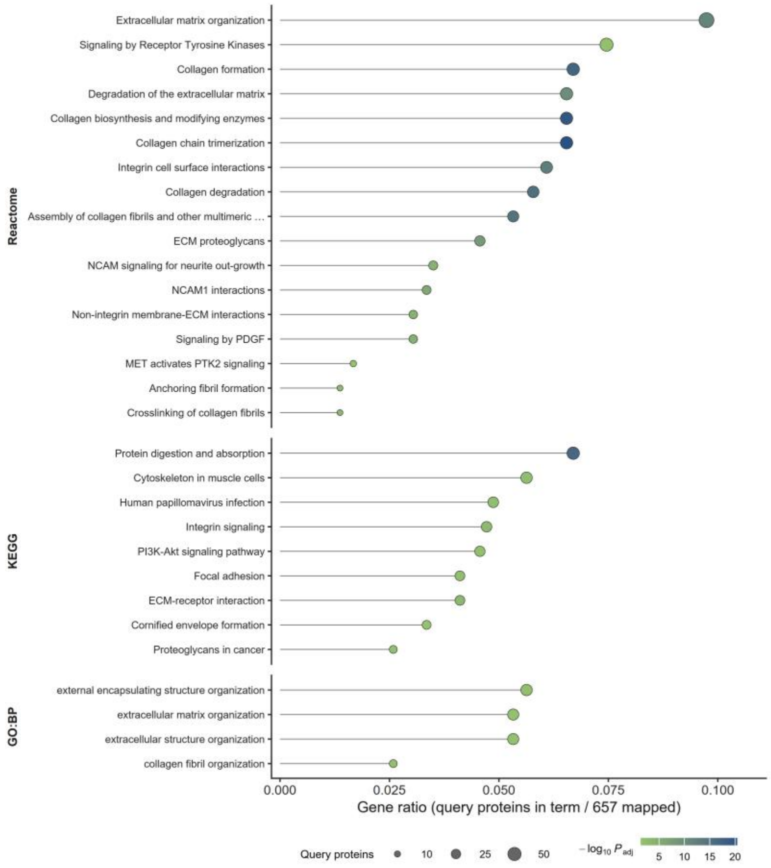


**Supplementary Figure S3 | Pathway over-representation of age-clock peptide proteins.**

The 30 specific terms reaching significance in a g:Profiler over-representation analysis of the parent proteins of the retained peptides, against a custom background of 2,448 proteins detectable in this CE–MS assay: 17 Reactome, 9 KEGG, and 4 GO Biological Process terms. Seven further significant GO Biological Process terms sit at the top of the ontology hierarchy (each annotating 280–1,056 background proteins, against 118 for the next largest term of any database) and are omitted from the figure as uninformative; all 37 significant terms are listed in Supplementary Table S5. Of the 685 parent proteins of the 2,631 retained peptides, 667 carried a gene symbol and 657 mapped to the g:Profiler namespace; gene ratios are expressed against those 657. Point position is the gene ratio (query proteins annotated to the term / 657), point area is the number of query proteins in the term, and colour is the −log10 Benjamini–Hochberg false-discovery-rate-adjusted P (significance threshold 0·05, custom domain scope). The same analysis populates Supplementary Table S5. CE–MS, capillary electrophoresis–mass spectrometry; GO, Gene Ontology; KEGG, Kyoto Encyclopedia of Genes and Genomes.

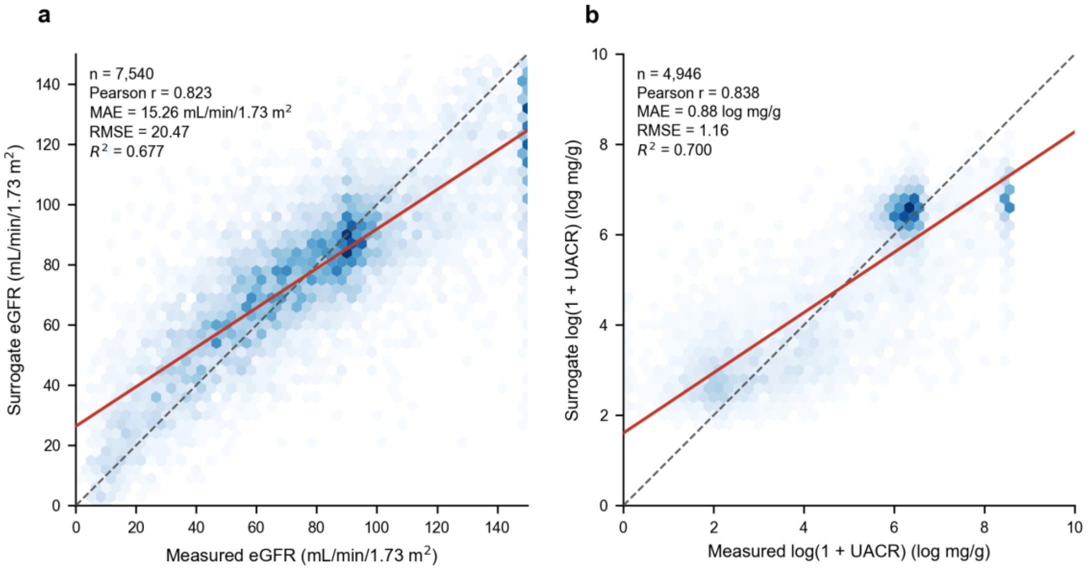


**Supplementary Figure S4 | Kidney-surrogate model accuracy.**

Out-of-fold performance of the two peptide-based kidney surrogates in the kidney-diverse cohort. Under five-fold cross-validation the eGFR surrogate reached Pearson r = 0·823 (MAE 15·26 mL/min/1·73 m²; RMSE 20·47; $R^2$ = 0·677; n = 7,540) and the UACR surrogate r = 0·838 (MAE 0·88; RMSE 1·16; $R^2$ = 0·700; n = 4,946). a,

Surrogate versus measured eGFR. b, Surrogate versus measured log-transformed UACR. Hexagonal bins show participant density; the dashed line is identity and the solid line the ordinary least-squares fit. Creatinine-based eGFR was capped at 150 mL/min/1·73 m² before surrogate modelling, as described in Supplementary Methods S4, which produces the density at the upper limit in panel a (409 observations: 408 capped from higher values and one already at 150). Both surrogates regress towards the mean at the extremes, which is why measured values are used wherever available and the surrogates serve only as the fallback arm of the dual-arm calibration. eGFR, estimated glomerular filtration rate; MAE, mean absolute error; RMSE, root mean squared error; UACR, urinary albumin-to-creatinine ratio.

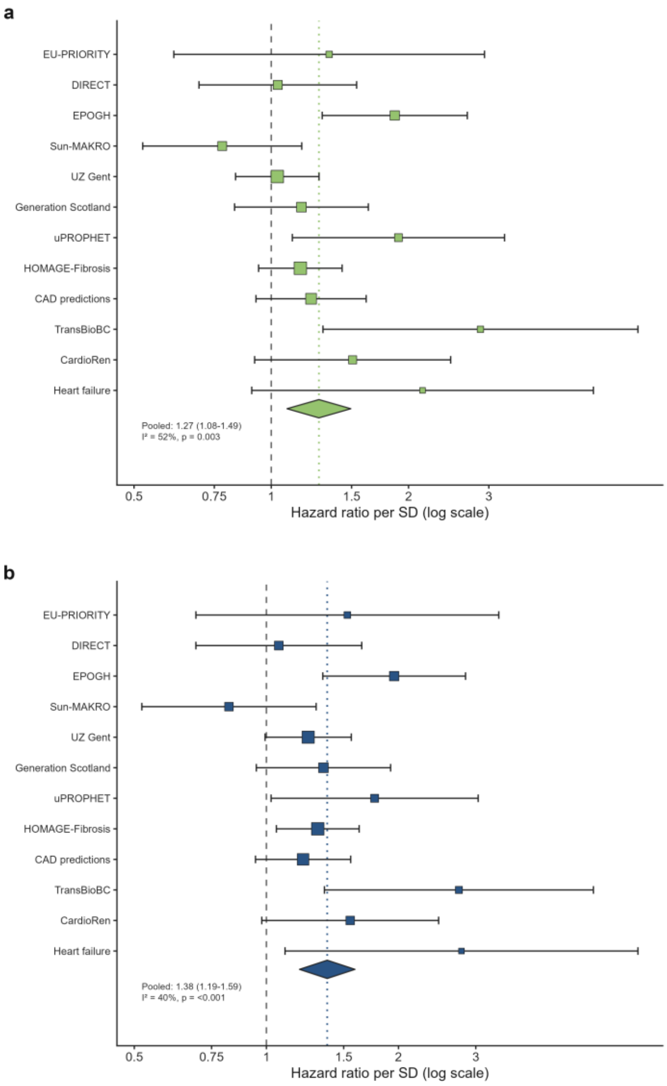


**Supplementary Figure S5 | Per-study meta-analysis of the association of UPBioAgeAcc and k-UPBioAgeAcc with all-cause mortality.**

Random-effects (DerSimonian–Laird) pooled hazard ratio per SD increment in a, UPBioAgeAcc (pooled HR 1·27, 95% CI 1·08–1·49; $I^2$ = 52%) and b, k-UPBioAgeAcc (pooled HR 1·38, 95% CI 1·19–1·59; $I^2$ = 39·8%). Models are fully adjusted for age, sex, diabetes, cardiovascular disease, hypertension, body-mass index, estimated glomerular filtration rate, and mean arterial pressure. Marker area is the inverse-variance weight, horizontal bars are 95% confidence intervals, and the diamond is the pooled estimate. Studies contributing fewer than 10 deaths (see Supplementary Table S1) were not entered individually into the per-study random-effects meta-analysis; where only one participant of a study differed on a yes/no covariate, that covariate was omitted from the study's model (cardiovascular disease in HOMAGE-Fibrosis, for both measures). HR, hazard ratio; SD, standard deviation; UPBioAgeAcc, urinary peptidomic age acceleration; k-UPBioAgeAcc, kidney-conditioned urinary peptidomic age acceleration.

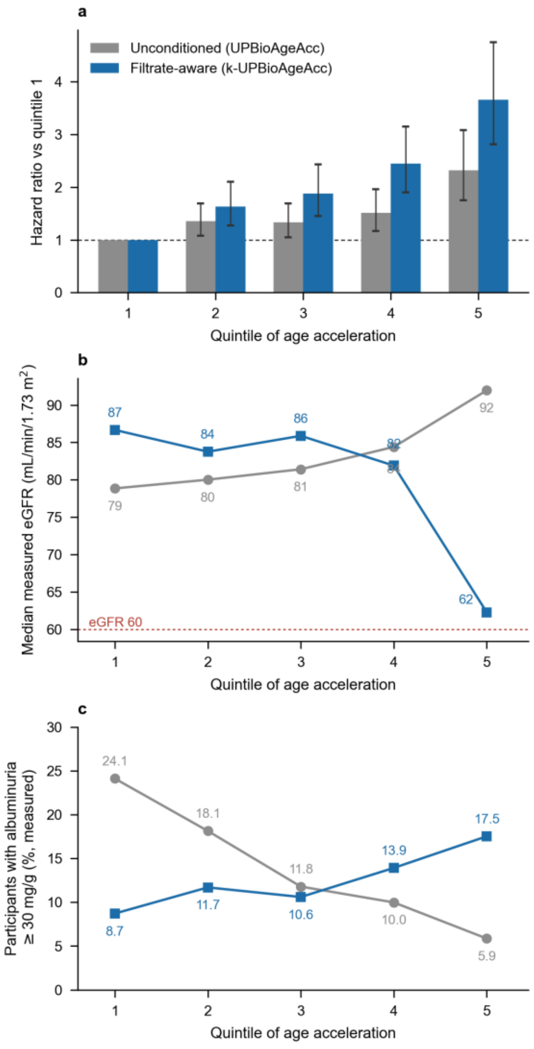


**Supplementary Figure S6 | Filtrate-Aware Calibration reverses a biologically implausible ordering.** Participants in the clinical follow-up cohort (n = 7,469; 625 deaths) were ranked into quintiles of unconditioned urinary age acceleration (UPBioAgeAcc) and, separately, of kidney-conditioned urinary age acceleration (k-UPBioAgeAcc). a, Hazard ratio for all-cause mortality relative to the lowest quintile, from Cox models adjusted for age and sex; bar heights are the hazard ratio and error bars the 95% confidence interval. b, Median measured eGFR of the same quintiles (dashed line, 60 mL/min/1·73 m²). c, Percentage of each quintile with a measured urinary albumin-to-creatinine ratio of at least 30 mg/g; panel c uses measured values only, and the number of participants with a measured ratio per quintile is 340 to 717 for the unconditioned measure and 445 to 546 for the kidney-conditioned measure. Before conditioning, median eGFR rises across quintiles of apparent biological age (79 to 92 mL/min/1·73 m²) while albuminuria falls (24·1% to 5·9%), so the participants ranked most aged are those with the best-preserved filtration; after conditioning both orderings reverse (87 to 62 mL/min/1·73 m²; 8·7% to 17·5%). Quintile boundaries were −3·62, +1·41, +5·93, and +11·52 years for the kidney-conditioned measure and −5·95, −1·21, +2·86, and +7·77 years for the unconditioned measure. eGFR, estimated glomerular filtration rate.

### Supplementary Tables

**Supplementary Table S1 | Per-study composition of the clinical follow-up cohort.**

| Study | No. Participants | No. Deaths | Mean (SD) age, yr | % Female | Sampling years | Ref. |
|---|---|---|---|---|---|---|
| EU-PRIORITY | 1,765 | 13 | 61·6 (8·3) | 37·8% | 2014–2016 | 31 |
| DIRECT | 1,486 | 39 | 45·3 (15·1) | 49·8% | 2001–2004 | 32 |
| EPOGH | 914 | 64 | 50·1 (15·4) | 49·5% | 2005–2015 | 10 |
| Sun-MACRO | 581 | 25 | 63·0 (9·2) | 21·0% | 2014 | 33 |
| UZ Gent | 488 | 117 | 62·0 (16·7) | 41·6% | 2011–2012 | 34 |
| Generation Scotland | 473 | 56 | 65·8 (12·5) | 32·6% | 2006–2011 | 35 |
| uPROPHET | 462 | 18 | 57·2 (14·8) | 25·5% | 2014–2016 | 36 |
| HOMAGE fibrosis | 354 | 125 | 70·2 (10·1) | 27·4% | 2010–2014 | 37 |
| PersTIgAN | 270 | 5 | 45·1 (14·3) | 34·8% | 2000–2019 | 38 |
| CAD Predictions | 145 | 95 | 70·9 (11·2) | 37·2% | 1999–2000 | 39 |
| TransBioBC | 131 | 14 | 65·2 (12·5) | 19·8% | 2007–2012 | 40 |
| CardioRen | 116 | 29 | 69·9 (9·7) | 26·7% | 2010–2012 | 41 |
| STOP-IgAN | 109 | 2 | 44·6 (12·7) | 23·9% | 2008–2011 | 42 |
| Heart failure | 84 | 17 | 66·0 (9·0) | 52·4% | 2005–2011 | 43 |
| Predictions (HIB) | 53 | 0 | 58·5 (10·4) | 20·8% | 2003–2004 | 44 |
| Predictions (Prague) | 22 | 6 | 66·9 (6·5) | 31·8% | 2007–2008 | 44 |
| Predictions (Groningen) | 16 | 0 | 65·1 (9·5) | 43·8% | 2008 | 44 |
| **Total** | **7,469** | **625** | 57·0 (15·3) | 38·2% | 1999–2019 | |

Number of participants, deaths, mean (SD) age and % female contributions for each of the 17 studies making up the clinical follow-up cohort. Age is presented at the time of urine sampling. Sampling years give the range of urine-collection dates; a collection date was recorded for 7,389 of 7,469 participants (Generation Scotland 394 of 473, STOP-IgAN 108 of 109). Ethical approval and written informed consent were obtained in each original recruitment study; references can be found in the numbered citation describing each study's design and recruitment. The present analysis used fully anonymised data held in the Human Urinary Proteome Database; the Ethics Committee of Hannover Medical School, Germany, has waived the requirement for ethical approval for analyses of such fully anonymised data (No. 3116-2016). FLEMENGHO does not appear as a separate row: 775 FLEMENGHO participants who were also followed for vital status are counted within the EPOGH (714) and Heart failure (61) rows. Studies are named by their entry in the source database. Ethnicity was not recorded in any contributing study; most recruited at European centres, while some, including DIRECT, Sun-MACRO, and CAD Predictions, recruited internationally. Each contributing study was funded independently; funding sources are stated in the primary publications cited for each study. EPOGH, European Project on Genes in Hypertension; FLEMENGHO, Flemish Study on Environment, Genes and Health Outcomes; HOMAGE, Heart OMics in AGEing.

**Supplementary Table S2 | Incidence, follow-up, and number of adverse health outcomes in the clinical follow-up cohort.**

| Endpoint | n/N | Median (IQR) follow-up, yr | Mean (SD) age, yr | % Female |
|---|---|---|---|---|
| All-cause mortality | 625/7,469 | 3·95 (2·39–5·58) | 57·0 (15·3) | 38·2% |
| Incident CAD | 363/5,560 | 3·76 (2·23–5·80) | 60·6 (12·6) | 38·9% |
| Incident HF | 471/5,560 | 3·76 (2·23–5·80) | 60·6 (12·6) | 38·9% |
| Composite CVD endpoint | 834/5,560 | 3·76 (2·23–5·80) | 60·6 (12·6) | 38·9% |
| CKD progression | 113/3,617 | 3·01 (2·12–4·37) | 59·0 (10·5) | 40·2% |

n = events, N = participants at risk; follow-up is median (IQR). Total person-time was 33,651 person-years for all-cause mortality, 25,225 person-years for the three cardiovascular endpoints, which are analysed on a single common risk set and therefore share one total, and 11,179 person-years for CKD progression. The corresponding crude rates per 1,000 person-years were 18·6 for all-cause mortality, 14·4 for incident coronary artery disease, 18·7 for incident heart failure, 33·1 for the composite cardiovascular endpoint, and 10·1 for CKD progression. CKD, chronic kidney disease.

**Supplementary Table S3 | Clinical follow-up cohort demographic characteristics.**

| Characteristic | Overall (n = 7,469) | Survived (n = 6,844) | Died (n = 625) |
|---|---|---|---|
| Number in group (%) | 7,469 (100%) | 6,844 (100%) | 625 (100%) |
| Age [yr] | 57·0 (15·3) | 55·7 (14·9) | 71·7 (10·7) |
| Female, n (%) | 2,854 (38·2%) | 2,655 (38·8%) | 199 (31·8%) |
| eGFR [mL/min/1·73 m²] | 78·8 (28·7) | 80·5 (28·2) | 61·0 (27·5) |
| Kidney disease, n (%) | 2,212 (29·6%) | 1,898 (27·7%) | 314 (50·2%) |
| Diabetes, n (%) | 4,096 (54·8%) | 3,933 (57·5%) | 163 (26·1%) |
| Cardiovascular disease, n (%) | 1,357 (18·2%) | 983 (14·4%) | 374 (59·8%) |
| Hypertension, n (%) | 3,088 (41·3%) | 2,756 (40·3%) | 332 (53·1%) |
| Follow-up [yr] | 4·5 (3·2) | 4·6 (3·2) | 3·6 (2·6) |

Continuous variables are presented as a mean with (SD); categorical variables as n (%). Percentages are column percentages within each group. Diabetes is more common among survivors than among those who died, the reverse of the pattern for cardiovascular and kidney disease. This reflects the composition of the contributing studies rather than a protective effect: the two largest studies are almost entirely diabetic and have low mortality (n = 1,765, 99·9% with diabetes, 0·7% died; and n = 1,486, 100% with diabetes, 2·6% died), whereas the larger studies with the highest mortality (CAD Predictions, HOMAGE fibrosis, CardioRen, and UZ Gent; 24·0–65·5% died) had diabetes in 0–30·3% of participants. Study-stratified and study-adjusted analyses are reported in Supplementary Methods S7. Abbreviations: eGFR, estimated glomerular filtration rate (mL/min/1·73 m²); UACR, urinary albumin-to-creatinine ratio (mg/g).

**Supplementary Table S4 | Top 20 contributing peptides to UPBioAge.**

| Rank | Peptide ID | Amino-acid sequence | Parent Protein | Meta coefficient | Biological axis |
|---|---|---|---|---|---|
| 1 | e213211 | DPNMEANSHYGHNDDVR | PECAM1 | +0·634 | Vascular / endothelial |
| 2 | e014305 | PGLMGESSESFSASEDEG | PHACTR2 | +0·501 | Cytoskeletal / actin regulation |
| 3 | e212208 | PGMGNADDEQQEEGT | GTF3C4 | +0·499 | Nuclear – RNA Pol III |
| 4 | e212221 | KGEEGSDDDETENGP | CTR9 | +0·472 | Nuclear – transcription |
| 5 | e017518 | YSISSDNSDTTDSHATSTSAS | JADE2 | +0·423 | Nuclear – chromatin |
| 6 | e017129 | HGSGSSYSYSRGHYESGSGQ | HRNR | +0·421 | Epithelial / cornified envelope |
| 7 | e014287 | SGGYGGGYGGQSSMSGYDQ | HNRNPH1/HNRNPH2 | +0·404 | Nuclear – RNA processing |
| 8 | e213213 | QGYSDSEERESMPRDGE | ZEB2 | +0·371 | Nuclear – transcription |
| 9 | e014074 | EEAPSLRPAPPPISGGGY | FGB | +0·341 | Coagulation / acute-phase |
| 10 | e213495 | STETRSSSSESSHSSSSHSH | RPS6KA5 | +0·335 | Nuclear – signalling kinase |
| 11 | e020065 | DDILASPPRLPEPQPYPGAPHHS | COL18A1 | +0·322 | ECM – basement membrane |
| 12 | e013869 | EDAGAGGEDAGPGGEDVGPGG | GOLGA6L2 | +0·313 | Golgi |
| 13 | e014303 | GGSSNSGGGSGSQSGSSGSGSNG | DMKN | +0·311 | Epithelial / keratinocyte |
| 14 | e013475 | KSEEEKDSESHLAED | ANK2 | +0·305 | Cytoskeletal / membrane scaffold |
| 15 | e004567 | MAGSGMGQSME | BCL9L | +0·303 | Nuclear – Wnt/transcription |
| 16 | e207689 | CDDYRLCE | MGP | +0·296 | Calcification inhibitor (tentative) |
| 17 | e000786 | SNTEERL | SEMG1 | +0·282 | Seminal plasma |
| 18 | e007501 | TDDSNDYGENE | ZNF652 | +0·282 | Nuclear – transcription |
| 19 | e214130 | SPGPVGFPGDPGPPGEPGPAGQDGPP | COL5A1 | +0·280 | ECM – fibrillar collagen |
| 20 | e020604 | FQTYGAIQGPPGQKGEMGTPGPKGD | COL17A1 | −0·278 | ECM – hemidesmosome |

Peptides are ranked by the absolute value of their Stage-2 elastic-net meta coefficient; the signed coefficient is given for each. Coefficients are in years of predicted age per standard deviation of that peptide's Stage-1 age signal, so a positive coefficient indicates that a higher peptide contribution raises the predicted age. Amino-acid sequences and parent-protein assignments are from the MS/MS annotation. The rank-7 sequence occurs identically in HNRNPH1 and HNRNPH2, so its source cannot be assigned to either; it is one model feature and is counted once among the 685 parent proteins. Biological axis provides an interpretive grouping. The share of model weight for all 2,631 retained peptides is in Supplementary Data 1, and the complete fitted model is in the Zenodo deposit (doi:10.5281/zenodo.22914811). UPBioAge, urinary peptidomic biological age.

**Supplementary Table S5 | Pathway over-representation of the age-clock peptide proteins.**

| Pathway ID | Pathway descriptor | q value | Key proteins |
|---|---|---|---|
| R-hsa-8948216 | Collagen chain trimerization | 4·03E−21 | COL1A1, COL1A2, COL3A1, COL4A1, COL4A3, COL5A2, COL7A1, COL17A1, COL25A1… |
| R-hsa-1650814 | Collagen biosynthesis and modifying enzymes | 3·40E−20 | COL10A1, COL11A1, COL11A2, COL12A1, COL13A1, COL14A1… |

| Pathway ID | Pathway descriptor | q value | Key proteins |
|---|---|---|---|
| R-hsa-1474290 | Collagen formation | 4·71E−18 | COL10A1, COL11A1, COL11A2, COL12A1, COL13A1, COL14A1… |
| R-hsa-1442490 | Collagen degradation | 9·75E−16 | COL1A1, COL1A2, COL3A1, COL4A1, COL4A3, COL5A2, COL7A1, COL17A1, COL25A1… |
| R-hsa-2022090 | Assembly of collagen fibrils and other multimeric structures | 4·59E−15 | COL1A1, COL2A1, COL3A1, COL5A2, COL10A1, COL11A1, COL14A1, COL17A1, COL18A1… |
| R-hsa-216083 | Integrin cell surface interactions | 1·87E−13 | COL1A1, COL1A2, COL4A1, COL4A3, COL5A2, FGA, PECAM1… |
| R-hsa-1474244 | Extracellular matrix organization | 2·93E−12 | COL1A1, COL1A2, COL3A1, COL4A1, FGA, COL5A2, COL7A1, COL17A1… |
| R-hsa-1474228 | Degradation of the extracellular matrix | 1·13E−10 | BCAN, CDH1, COL10A1, COL11A1, COL11A2, COL12A1… |
| R-hsa-3000178 | ECM proteoglycans | 9·75E−09 | AGRN, BCAN, COL1A1, COL1A2, COL2A1, COL3A1… |
| R-hsa-419037 | NCAM1 interactions | 1·75E−06 | COL2A1, COL3A1, COL4A1, COL4A2, COL4A3, COL4A4, COL4A5, COL5A1, COL5A2… |
| R-hsa-186797 | Signaling by PDGF | 4·12E−05 | COL2A1, COL3A1, COL4A1, COL4A2, COL4A3, COL4A4… |
| R-hsa-375165 | NCAM signaling for neurite out-growth | 6·87E−05 | AGRN, CACNA1C, CACNA1D, CACNA1H, COL2A1, COL3A1… |
| R-hsa-3000171 | Non-integrin membrane-ECM interactions | 9·16E−05 | AGRN, COL10A1, COL11A1, COL11A2, COL1A1, COL1A2… |
| R-hsa-2243919 | Crosslinking of collagen fibrils | 6·22E−04 | COL1A1, COL1A2, COL4A1, COL4A2, COL4A3, COL4A4… |
| R-hsa-2214320 | Anchoring fibril formation | 4·42E−03 | COL1A1, COL1A2, COL4A1, COL4A2, COL4A3, COL4A4… |
| R-hsa-9006934 | Signaling by Receptor Tyrosine Kinases | 2·19E−02 | ACTB, ALK, APOE, BRAF, CDK5R2, COL11A1… |
| R-hsa-8874081 | MET activates PTK2 signaling | 3·95E−02 | COL11A1, COL11A2, COL1A1, COL1A2, COL24A1, COL27A1… |
| KEGG:04974 | Protein digestion and absorption | 1·92E−17 | COL10A1, COL11A1, COL11A2, COL12A1, COL13A1, COL14A1… |
| KEGG:04512 | ECM-receptor interaction | 1·05E−03 | AGRN, COL1A1, COL1A2, COL2A1, COL4A1, COL4A2… |
| KEGG:04518 | Integrin signaling | 1·12E−03 | ACTB, C3, CDH1, COL1A1, COL1A2, COL2A1… |
| KEGG:04820 | Cytoskeleton in muscle cells | 8·07E−03 | ACTB, AGRN, ANK2, COL11A1, COL11A2, COL1A1… |
| KEGG:05165 | Human papillomavirus infection | 8·87E−03 | APC, COL1A1, COL1A2, COL2A1, COL4A1, COL4A2… |
| KEGG:04510 | Focal adhesion | 1·37E−02 | ACTB, BRAF, COL1A1, COL1A2, COL2A1, COL4A1… |
| KEGG:04151 | PI3K-Akt signaling pathway | 1·53E−02 | COL1A1, COL1A2, COL2A1, COL4A1, COL4A2, COL4A3… |
| KEGG:05205 | Proteoglycans in cancer | 1·69E−02 | ACTB, ANK2, ARHGEF1, ARHGEF12, BRAF, COL1A1… |
| KEGG:04382 | Cornified envelope formation | 3·55E−02 | COL17A1, COL4A1, COL4A2, COL4A3, COL4A4, COL4A5… |
| GO:0009888 | Tissue development | 4·97E−03 | ACTB, ADM, AKAP6, AMTN, ANKH, APC… |
| GO:0048513 | Animal organ development | 6·55E−03 | ACTB, ADM, AFF2, AKAP6, ALK, AMTN… |
| GO:0050793 | Regulation of developmental process | 6·55E−03 | ABCA1, ACTB, ADGRA2, ADM, ADRA2C, AGRN… |
| GO:0045229 | External encapsulating structure organization | 1·05E−02 | COL11A1, COL11A2, COL12A1, COL14A1, COL19A1, COL1A1… |
| GO:0030199 | Collagen fibril organization | 1·05E−02 | COL11A1, COL11A2, COL12A1, COL14A1, COL1A1, COL1A2… |
| GO:0032502 | Developmental process | 2·47E−02 | ABCA1, ACRV1, ACTB, ACTL9, ADGRA2, ADM… |
| GO:0043062 | Extracellular structure organization | 2·51E−02 | COL11A1, COL11A2, COL12A1, COL14A1, COL19A1, COL1A1… |
| GO:0030198 | Extracellular matrix organization | 2·51E−02 | COL11A1, COL11A2, COL12A1, COL14A1, COL19A1, COL1A1… |
| GO:0051253 | Negative regulation of RNA metabolic process | 4·30E−02 | BASP1, BCOR, BMAL1, CC2D1B, CCAR1, CDX4… |

| Pathway ID | Pathway descriptor | q value | Key proteins |
|---|---|---|---|
| GO:0030154 | Cell differentiation | 4·41E−02 | ABCA1, ACTB, ACTL9, ADGRA2, ADM, ADRA2C… |
| GO:0048869 | Cellular developmental process | 4·48E−02 | ABCA1, ACTB, ACTL9, ADGRA2, ADM, ADRA2C… |

Over-representation analysis of the parent proteins of the 2,631 retained peptides, run with g:Profiler against a custom background of the CE–MS-detectable urinary proteome (effective domain 2,448 proteins) rather than the whole genome, so enrichment is judged against what this assay can observe. Terms are Benjamini–Hochberg significant at FDR 0·05 and are listed by ascending q value within each source; the key proteins column gives the query proteins driving each term and is truncated where a term carries many. Because the urinary peptidome consists of proteolytic fragments, enrichment of collagen synthesis, trimerisation, and modification terms indexes collagen turnover and matrix homeostasis rather than net synthesis. The query proteins are the parent proteins of model peptides rather than an independently selected gene set, so this analysis is descriptive: it characterises what the clock is built from, and no causal or mechanistic claim follows from it. FDR, false discovery rate.

**Supplementary Table S6 | Chronological-age prediction performance of the k-UPBioAge clock across analysis cohorts.**

| Cohorts | n | Age, yr | k-UPBioAge, yr | MAE (95% CI), yr | RMSE, yr | Pearson r (95% CI) | $R^2$ |
|---|---|---|---|---|---|---|---|
| *Derivation cohort* | | | | | | | |
| Train (out-of-fold) | 1,449 | 48·5 (17·9) | 48·6 (19·3) | 5·91 (5·68–6·16) | 7·59 | 0·92 (0·91–0·93) | 0·82 |
| Test | 362 | 48·4 (18·0) | 48·9 (19·3) | 4·94 (4·54–5·36) | 6·37 | 0·94 (0·93–0·95) | 0·87 |
| *Validation cohorts* | | | | | | | |
| Clinical follow-up cohort | 7,469 | 57·0 (15·3) | 61·1 (14·9) | 7·93 (7·79–8·08) | 10·21 | 0·81 (0·80–0·82) | 0·55 |
| Glasgow cohort | 675 | 26·3 (6·4) | 30·2 (8·7) | 5·44 (5·10–5·76) | 6·97 | 0·74 (0·71–0·77) | −0·17 |
| FLEMENGHO | 778 | 51·0 (15·8) | 51·1 (15·0) | 5·44 (5·11–5·77) | 7·15 | 0·89 (0·88–0·91) | 0·80 |

Data for age and k-UPBioAge presented as a mean with (SD). 95% CIs are from a participant-level bootstrap (2,000 resamples) for the mean absolute error and from the Fisher z transformation for Pearson r. The derivation training row is an internal out-of-fold calibration metric, not an independent validation result, because the peptide screen was performed once in the whole training set; the held-out test row and the validation cohorts carry the prediction-accuracy claim. All errors reported here are calibrated rather than age-blind, because the clock’s age-bias correction is evaluated at each participant’s own chronological age; the corresponding errors before that correction are given in Supplementary Methods S3. All performance metrics are for the kidney-conditioned clock (k-UPBioAge); corresponding values for the unconditioned UPBioAge clock are given in Supplementary Methods S3. Abbreviations: FLEMENGHO: Flemish Study on Environment, Genes and Health Outcomes; MAE: mean absolute error; RMSE: root-mean-squared error; $R^2$: coefficient of determination. $R^2$ is negative in the Glasgow cohort because squared prediction error there exceeds the error of predicting that cohort’s own mean age; this is compatible with r = 0·74 and reflects the narrow age range (SD 6·4 years) together with a mean prediction 3·8 years above mean chronological age. That cohort supports transferability of the age-associated signal rather than of absolute age calibration. UPBioAge, urinary peptidomic biological age; k-UPBioAge, kidney-conditioned urinary peptidomic biological age.

**Supplementary Table S7 | Prediction accuracy of k-UPBioAge, and the effect of kidney conditioning on age acceleration, by cohort and by clinical stratum.**

| Cohort | n | MAE (yr) | RMSE (yr) | r | $R^2$ | Mean (SD) UPBioAgeAcc (yr) | Mean (SD) k-UPBioAgeAcc (yr) | Cohen's dz (paired) |
|---|---|---|---|---|---|---|---|---|
| Glasgow | 675 | 5·44 | 6·97 | 0·741 | −0·17* | 3·22 (5·84) | 3·53 (5·82) | 0·224 |
| FLEMENGHO | 778 | 5·44 | 7·15 | 0·894 | 0·796 | −0·36 (7·04) | −0·03 (7·10) | 0·220 |
| Clinical follow-up cohort | 7,469 | 7·93 | 10·21 | 0·808 | 0·554 | 0·87 (8·52) | 4·05 (9·32) | 0·756 |
| No major comorbidity | 690 | 5·83 | 7·60 | 0·873 | 0·755 | −0·40 (7·52) | 0·37 (7·55) | 0·353 |
| CVD only | 343 | 6·70 | 9·19 | 0·824 | 0·663 | −2·70 (8·31) | 0·53 (9·13) | 0·875 |
| Diabetes only | 2,251 | 7·42 | 9·18 | 0·840 | 0·641 | 3·14 (8·32) | 3·79 (8·24) | 0·350 |
| Hypertension only | 611 | 6·38 | 8·20 | 0·816 | 0·632 | −2·35 (7·67) | −1·02 (8·11) | 0·479 |
| Kidney disease only | 434 | 11·46 | 14·49 | 0·806 | 0·361 | 3·07 (9·62) | 9·45 (10·85) | 1·498 |
| Multiple comorbidities | 3,140 | 8·71 | 11·12 | 0·677 | 0·091 | 0·23 (8·39) | 5·66 (9·55) | 1·213 |
| eGFR G1 (≥90) | 2,937 | 7·33 | 9·24 | 0·823 | 0·633 | 2·35 (8·58) | 3·08 (8·60) | 0·361 |
| eGFR G2 (60–89) | 2,627 | 6·68 | 8·73 | 0·762 | 0·507 | −0·89 (7·57) | 1·80 (8·50) | 0·709 |
| eGFR G3a (45–59) | 732 | 9·35 | 11·87 | 0·668 | 0·145 | 0·10 (8·77) | 6·57 (9·85) | 1·706 |
| eGFR G3b (30–44) | 700 | 10·38 | 12·81 | 0·726 | 0·055 | 0·64 (8·72) | 8·93 (9·15) | 2·228 |
| eGFR G4 (15–29) | 433 | 12·36 | 14·73 | 0·648 | −0·457 | 2·48 (9·96) | 10·95 (9·86) | 2·702 |
| eGFR G5 (<15) | 40 | 17·56 | 20·18 | 0·744 | −0·240 | 7·33 (12·15) | 15·75 (12·72) | 3·497 |
| Albuminuria A1 (<30) | 2,173 | 7·28 | 9·14 | 0·837 | 0·645 | 3·56 (8·27) | 3·49 (8·32) | −0·146 |
| Albuminuria A2 (30–299) | 217 | 7·33 | 9·36 | 0·871 | 0·684 | −0·65 (8·20) | 4·27 (8·30) | 1·616 |
| Albuminuria A3 (≥300) | 92 | 11·12 | 13·35 | 0·811 | 0·244 | −1·75 (9·05) | 9·86 (8·99) | 6·680 |

All performance metrics are for the kidney-conditioned prediction (k-UPBioAge). Cohen's dz is the paired effect size of k-UPBioAgeAcc against UPBioAgeAcc within the same participants, mean(difference)/SD(difference); it measures how far kidney conditioning moves age acceleration in that stratum, not a comparison with any other cohort. *$R^2$ is computed against each cohort's own age variance. The negative value in the Glasgow cohort is explained in the Supplementary Table S6 note. The final nine rows stratify the clinical follow-up cohort by KDIGO eGFR category and by albuminuria category. Denominators differ between the two blocks: eGFR was measured in all 7,469 participants, whereas the albuminuria rows cover only the 2,482 with a measured UACR. Accuracy is poorest at the lowest filtration (mean absolute error 17·56 years in G5, against 7·33 years in G1 and 6·68 years in G2; the unconditioned prediction gives 11·51 years in G5 and 6·00 years in G2), and the negative $R^2$ values in G4 and G5 should be interpreted cautiously, since $R^2$ is computed against the age variance within each stratum and that variance is narrow in these small groups. Age acceleration widens across eGFR categories after conditioning rather than before it (a spread of 13·9 years for k-UPBioAgeAcc against 8·2 years for UPBioAgeAcc), which is the expected direction if impaired filtration suppresses the peptide signal on which the clock depends and conditioning offsets that suppression. The residual in advanced chronic kidney disease is consistent with accelerated ageing, although the present data cannot separate that from a residual filtration artefact. eGFR, estimated glomerular filtration rate; KDIGO, Kidney Disease: Improving Global Outcomes; MAE, mean absolute error; RMSE, root mean squared error; SD, standard deviation; UACR, urinary albumin-to-creatinine ratio; UPBioAgeAcc, urinary peptidomic age acceleration; k-UPBioAgeAcc, kidney-conditioned urinary peptidomic age acceleration.

**Supplementary Table S8 | Risk for adverse outcomes in association with kidney-conditioned urinary peptidomic age acceleration (k-UPBioAgeAcc).**

| | n/N (%) | HR (95% CI) | p value |
|---|---|---|---|
| Mortality | | | |
| Total | 625/7,469 (8·4%) | | |
| Basic model | | 1·62 (1·49–1·76) | <0·0001 |
| Fully adjusted model | | 1·47 (1·34–1·62) | <0·0001 |
| Secondary incident endpoints | | | |
| Composite CVD endpoint | 834/5,560 (15·0%) | | |
| Basic model | | 1·68 (1·56–1·81) | <0·0001 |
| Fully adjusted model | | 1·38 (1·27–1·50) | <0·0001 |
| Heart failure | 471/5,560 (8·5%) | | |
| Basic model | | 1·75 (1·59–1·93) | <0·0001 |
| Fully adjusted model | | 1·29 (1·15–1·44) | <0·0001 |
| CAD | 363/5,560 (6·5%) | | |
| Basic model | | 1·60 (1·43–1·79) | <0·0001 |
| Fully adjusted model | | 1·44 (1·27–1·64) | <0·0001 |
| CKD progression | 113/3,617 (3·1%) | | |
| Basic model | | 2·66 (2·13–3·32) | <0·0001 |
| Fully adjusted model | | 1·39 (1·08–1·78) | 0·010 |

Hazard ratios per 1 SD of k-UPBioAgeAcc in the clinical follow-up cohort, by Cox regression using Efron's method for ties, without penalisation (95% CIs are Wald). Each endpoint block reports events over participants at risk, under the basic model (age and sex) and the fully adjusted model (age, sex, body-mass index, measured eGFR, mean arterial pressure, diabetes, cardiovascular disease, and hypertension). SD, standard deviation.

**Supplementary Table S9 | Mortality association of k-UPBioAgeAcc within strata of the clinical follow-up cohort, by CKD stage, albuminuria category, age group, sex, and comorbidity.**

| Stratum | n/N (%) | HR (95% CI) | p value |
|---|---|---|---|
| **CKD stage (interaction p=0·031)** | | | |
| G5 (eGFR <15) | 14/40 (35·0%) | 1·62 (0·60–4·36) | 0·34 |
| G4 (eGFR 15–29) | 84/433 (19·4%) | 0·93 (0·71–1·22) | 0·61 |
| G3b (eGFR 30–44) | 102/700 (14·6%) | 1·36 (1·07–1·74) | 0·013 |
| G3a (eGFR 45–59) | 105/732 (14·3%) | 1·41 (1·10–1·82) | 0·0072 |
| G2 (eGFR 60–89) | 224/2,627 (8·5%) | 1·64 (1·42–1·89) | $6{\cdot}9\times10^{-12}$ |
| G1 (eGFR ≥90) | 96/2,937 (3·3%) | 1·62 (1·30–2·02) | $1{\cdot}6\times10^{-5}$ |
| **Albuminuria category, measured UACR only (interaction p=0·057)** | | | |
| A1 (<30 mg/g) | 91/2,173 (4·2%) | 1·32 (1·01–1·72) | 0·040 |
| A2 (30–299 mg/g) | 75/217 (34·6%) | 1·43 (1·10–1·86) | 0·0069 |
| A3 (≥300 mg/g) | 34/92 (37·0%) | 0·86 (0·56–1·31) | 0·48 |
| **Age group (interaction p=0·27)** | | | |
| 18–<60 yr | 78/3,719 (2·1%) | 1·54 (1·18–2·02) | 0·0014 |
| 60–<70 yr | 145/2,196 (6·6%) | 1·52 (1·26–1·85) | $1{\cdot}9\times10^{-5}$ |
| 70–<80 yr | 251/1,230 (20·4%) | 1·57 (1·33–1·84) | $4{\cdot}0\times10^{-8}$ |
| ≥80 yr | 151/318 (47·5%) | 1·34 (1·10–1·62) | 0·0030 |
| **Sex (interaction p=0·53)** | | | |
| Female | 199/2,854 (7·0%) | 1·50 (1·26–1·79) | $5{\cdot}3\times10^{-6}$ |
| Male | 426/4,615 (9·2%) | 1·45 (1·30–1·63) | $8{\cdot}9\times10^{-11}$ |
| **Comorbidity** | | | |
| **No diagnosed disease** | 25/690 (3·6%) | 1·76 (1·07–2·89) | 0·026 |
| **Diabetes (interaction p=0·98)** | | | |
| Present | 163/4,096 (4·0%) | 1·47 (1·20–1·79) | $2{\cdot}1\times10^{-4}$ |
| Absent | 462/3,373 (13·7%) | 1·61 (1·44–1·79) | $6{\cdot}9\times10^{-18}$ |
| **Kidney disease (interaction p=0·21)** | | | |
| Present | 314/2,212 (14·2%) | 1·37 (1·21–1·56) | $1{\cdot}2\times10^{-6}$ |
| Absent | 311/5,257 (5·9%) | 1·58 (1·38–1·81) | $2{\cdot}7\times10^{-11}$ |

| Stratum | n/N (%) | HR (95% CI) | p value |
|---|---|---|---|
| **Hypertension (interaction p=0·71)** | | | |
| Present | 332/3,088 (10·8%) | 1·51 (1·32–1·72) | $8{\cdot}2\times10^{-10}$ |
| Absent | 293/4,381 (6·7%) | 1·41 (1·23–1·62) | $8{\cdot}2\times10^{-7}$ |
| **Cardiovascular disease (interaction p=0·59)** | | | |
| Present | 374/1,357 (27·6%) | 1·51 (1·34–1·71) | $1{\cdot}8\times10^{-11}$ |
| Absent | 251/6,112 (4·1%) | 1·35 (1·16–1·58) | $1{\cdot}2\times10^{-4}$ |

HR (95% CI) for all-cause mortality per SD increment in k-UPBioAgeAcc; n = deaths and N = participants at risk. Hazard ratios for the CKD stage and albuminuria rows are per standard deviation of k-UPBioAgeAcc computed within each stratum; all other rows are per standard deviation computed across the whole clinical follow-up cohort (9·3 years). The two scales are not interchangeable, so hazard ratios should not be compared directly between these blocks. All rows use the fully adjusted model of the primary analysis (age, sex, diabetes, cardiovascular disease, hypertension, body-mass index, eGFR, and mean arterial pressure). Where a covariate defines the stratum it is also omitted, because adjusting for it would remove the contrast being estimated: the CKD stage, albuminuria, and kidney disease rows all omit eGFR (a kidney disease diagnosis rests largely on eGFR). Three rows need one further omission because cardiovascular disease is too closely tied to death within them for the model to converge: the G1 (eGFR ≥ 90) row and the two diabetes rows also omit it. Interaction P values for the four comorbidity strata are from a multiplicative k-UPBioAgeAcc × stratum term fitted in the whole cohort with adjustment for age, sex, and the four comorbidity indicators; for the CKD stage and albuminuria strata the interaction term is fitted with the same covariates as the rows beneath it. Benjamini–Hochberg correction was applied to the four comorbidity interaction tests (diabetes, cardiovascular disease, hypertension, and kidney disease), which are the multiplicity family defined in Supplementary Methods S7; the P values shown for these four strata are unadjusted, and after correction they are 0·86 for kidney disease, 0·95 for cardiovascular disease and hypertension, and 0·98 for diabetes. The CKD stage, albuminuria, age group, and sex rows are reported without correction. The age-group rows sum to 7,463 of the 7,469 participants in the clinical follow-up cohort; the remaining 6 were below 18 years of age (minimum 15·1 yr) and fall outside all four bands. CKD, chronic kidney disease; eGFR, estimated glomerular filtration rate; HR, hazard ratio; SD, standard deviation; k-UPBioAgeAcc, kidney-conditioned urinary peptidomic age acceleration.

**Supplementary Table S10 | eGFR and UACR attenuation of the k-UPBioAgeAcc mortality association.**

| | n/N (%) | Model 1 HR (95% CI) | Model 2 HR (95% CI) | %attenuation (95% CI)* |
|---|---|---|---|---|
| **eGFR attenuation** | 625/7,469 (8·4%) | 1·53 (1·40–1·66) | 1·46 (1·33–1·61) | 10·2 (0·3–19·3) |
| **UACR attenuation, measured albuminuria** | 200/2,482 (8·1%) | 1·79 (1·53–2·11) | 1·34 (1·12–1·61) | 50·0 (30·2–79·5) |
| **UACR attenuation, measured or estimated albuminuria** | 625/7,469 (8·4%) | 1·46 (1·33–1·61) | 1·27 (1·15–1·40) | 37·8 (25·3–51·7) |
| **UACR attenuation, estimated albuminuria** | 425/4,987 (8·5%) | 1·33 (1·19–1·50) | 1·21 (1·08–1·35) | 34·9 (21·4–58·8) |

HR (95% CI) for all-cause mortality per SD increment in k-UPBioAgeAcc. All rows use the same SD, that of k-UPBioAgeAcc in the full clinical follow-up cohort, so the hazard ratios are directly comparable between rows. Model 1 for the eGFR analysis: adjusted for age, sex, body-mass index, diabetes, cardiovascular disease, and hypertension. Model 1 for the three albuminuria analyses: adjusted for the same covariates plus measured eGFR, so that each analysis omits only the kidney measure whose attenuation it estimates. Model 2: additionally adjusted for eGFR or albuminuria in the eGFR and albuminuria analyses, respectively. The three albuminuria rows differ only in which participants contribute and which albuminuria value is used: a measured urinary albumin-to-creatinine ratio where one exists (2,482 participants, 33% of the cohort), the value actually used by the calibration in every participant, which is the measurement where one exists and a peptide-based estimate otherwise, and the peptide-based estimate in the 4,987 participants who have no measurement. In the measured-albuminuria subset only, prevalent cardiovascular disease is near-separating (190 participants carry 107 of the 200 deaths, an event rate of 56·3% against 4·1%), so that row is adjusted without it, as is the albuminuria block of Supplementary Table S14; retaining it gives a covariate hazard ratio of 4·1 and an attenuation of 31·3% (Model 1 HR 1·51, Model 2 HR 1·32). All other rows retain cardiovascular disease. *Percent reduction in the log hazard ratio per SD when the kidney measure is added to Model 1, with 95% CIs from participant-level bootstrap resampling. n = deaths and N = participants at risk. HR, hazard ratio; UACR, urinary albumin-to-creatinine ratio; k-UPBioAgeAcc, kidney-conditioned urinary peptidomic age acceleration.

**Supplementary Table S11 | Mortality and incident-disease associations of the kidney-conditioned and unconditioned urinary age acceleration, side by side.**

| Endpoint | Covariate adjustment | UPBioAgeAcc HR (95% CI) | k-UPBioAgeAcc HR (95% CI) | Ratio of hazard ratios (95% CI) | P |
|---|---|---|---|---|---|
| **All-cause mortality (n = 7,469; 625 events)** | | | | | |
| | Age and sex | 1·32 (1·21–1·45) | 1·62 (1·49–1·76) | 1·23 (1·17–1·28) | <0·001 |
| | Fully adjusted | 1·21 (1·11–1·33) | 1·47 (1·34–1·62) | 1·21 (1·16–1·27) | <0·001 |
| **Incident coronary artery disease (n = 5,560; 363 events)** | | | | | |
| | Age and sex | 1·17 (1·04–1·32) | 1·60 (1·43–1·79) | 1·36 (1·28–1·45) | <0·001 |
| | Fully adjusted | 1·13 (1·00–1·27) | 1·44 (1·27–1·64) | 1·28 (1·20–1·37) | <0·001 |
| **Incident heart failure (n = 5,560; 471 events)** | | | | | |
| | Age and sex | 1·34 (1·20–1·48) | 1·75 (1·59–1·93) | 1·31 (1·24–1·39) | <0·001 |
| | Fully adjusted | 1·08 (0·97–1·19) | 1·29 (1·15–1·44) | 1·20 (1·14–1·25) | <0·001 |
| **CKD progression (n = 3,617; 113 events)** | | | | | |
| | Age and sex | 1·41 (1·11–1·78) | 2·66 (2·13–3·32) | 1·89 (1·64–2·17) | <0·001 |
| | Fully adjusted | 1·03 (0·82–1·29) | 1·39 (1·08–1·78) | 1·35 (1·22–1·50) | 0·001 |
| **Composite cardiovascular endpoint (n = 5,560; 834 events)** | | | | | |
| | Age and sex | 1·26 (1·17–1·37) | 1·68 (1·56–1·81) | 1·33 (1·28–1·39) | <0·001 |
| | Fully adjusted | 1·11 (1·02–1·20) | 1·38 (1·27–1·50) | 1·25 (1·20–1·30) | <0·001 |

Hazard ratios are per 1-SD increment, from Cox proportional-hazards models fitted in the clinical follow-up cohort, with the same covariate sets applied to both measures so that each row is a like-for-like comparison. UPBioAgeAcc is the unconditioned urinary peptidomic age acceleration and k-UPBioAgeAcc the kidney-conditioned measure. The fully adjusted model is age, sex, diabetes, cardiovascular disease, hypertension, body-mass index, eGFR, and mean arterial pressure. The composite cardiovascular endpoint row above reproduces the k-UPBioAgeAcc estimates in Supplementary Table S8 exactly at both adjustment levels (Cox proportional-hazards models with Efron handling of ties, matching every other row in this table). The final two columns give the ratio of the two hazard ratios in that row (k-UPBioAgeAcc divided by UPBioAgeAcc), a value above 1 meaning the kidney-conditioned measure carries the stronger association in the same participants. Because the two estimates come from the same people they are correlated, so overlapping confidence intervals in the two hazard-ratio columns are not a valid test of the difference; the confidence interval and P value for the ratio instead come from a paired bootstrap over 2,000 resamples of participants, both models being refitted within each resample. P values are Benjamini–Hochberg-adjusted across the ten comparisons. The unconditioned measure did not perform better in any resample of any row, other than in one resample for CKD progression under full adjustment. For CKD progression under full adjustment, the endpoint with the fewest events, the interval rests on the 1,468 of 2,000 resamples in which both models converged. CKD, chronic kidney disease; eGFR, estimated glomerular filtration rate; UPBioAgeAcc, urinary peptidomic age acceleration; k-UPBioAgeAcc, kidney-conditioned urinary peptidomic age acceleration.

**Supplementary Table S12 | Discrimination for all-cause mortality, with and without the kidney-conditioned urinary age acceleration.**

| **Metric** | **Covariate ladder** | **n** | **Events** | **C-index baseline** | **C-index + metric** | **ΔC** | NRI (5-yr) | IDI (5-yr) |
|---|---|---|---|---|---|---|---|---|
| k-UPBioAgeAcc | Basic (age, sex) | 7,469 | 625 | 0·798 | 0·815 | +0·0179 (0·0094–0·0255) | 0·264 (0·164–0·368) | 0·033 (0·023–0·044) |
| k-UPBioAgeAcc | Fully adjusted | 7,469 | 625 | 0·830 | 0·835 | +0·0047 (0·0005–0·0089) | 0·20 (0·11–0·30) | 0·017 (0·010–0·026) |

C-index of the prognostic model with and without addition of the kidney-conditioned urinary peptidomic age acceleration (k-UPBioAgeAcc), in the clinical follow-up cohort (n = 7,469; 625 events), at the two levels of adjustment reported in the main text: Basic (age and sex) and the fully adjusted model (age, sex, body-mass index, measured eGFR, mean arterial pressure, diabetes, cardiovascular disease, and hypertension). ΔC = C-index gain from adding k-UPBioAgeAcc to the baseline model. All estimates in this table were computed out-of-fold: the cohort was divided into five parts and the division repeated five times, so that each participant's predicted risk came from a model that had not been fitted on them. The 95% confidence intervals shown come from 500 bootstrap resamples of the out-of-fold predictions, so they are conditional on the fitted cross-validation models. Because each fold's Cox model centres its relative hazard on its own training data, the gains were recomputed on the same folds with concordance counted only within each held-out fold; the point estimates were unchanged (fully adjusted ΔC 0·005, 95% CI 0·001–0·009; basic 0·018, 0·010–0·025, against the bootstrap intervals in the table). The net reclassification improvement (NRI) and the integrated discrimination improvement (IDI) are given at a five-year horizon. Both were computed in their continuous form on five-year predicted risks taken from the Cox models, with censoring handled by the Kaplan–Meier-based approach of Pencina and colleagues for censored time-to-event outcomes for the NRI, and by inverse-probability-of-censoring weighting for the IDI, so that participants censored before five years contribute.[12,13] The continuous NRI is the net difference in the proportions of events and non-events whose predicted risk moved in the correct direction; it is not the proportion of participants correctly reclassified. Model fit was compared in addition to discrimination. Adding k-UPBioAgeAcc to the Basic model gave a likelihood-ratio $\chi^2$ of 124·2 on 1 degree of freedom ($p=7{\cdot}7\times10^{-29}$) and lowered the Akaike information criterion from 9,312·4 to 9,190·3 (ΔAIC −122·2); adding it to the fully adjusted model gave $\chi^2$ = 64·2 on 1 degree of freedom ($p=1{\cdot}1\times10^{-15}$) and lowered the Akaike information criterion from 9,089·1 to 9,026·9 (ΔAIC −62·2). Differences in the Akaike information criterion were computed before rounding, so they do not equal the difference of the two rounded values printed here. Unlike the discrimination measures above, these two statistics come from models fitted in the whole cohort rather than out-of-fold.

**Supplementary Table S13 | Head-to-head comparison against the published UPP-age clock.**

| Cohort | Clock | MAE (yr) | RMSE (yr) | r | $R^2$ |
|---|---|---|---|---|---|
| Derivation test; n = 362 | k-UPBioAge | 4·94 | 6·37 | 0·944 | 0·874 |
| | UPP-age | 9·45 | 11·94 | 0·749 | 0·558 |
| Derivation train; n = 1,449 | k-UPBioAge | 5·91 | 7·59 | 0·920 | 0·821 |
| | UPP-age | 9·77 | 12·07 | 0·747 | 0·546 |
| Glasgow; n = 675 | k-UPBioAge | 5·44 | 6·97 | 0·741 | −0·17* |
| | UPP-age | 9·21 | 11·47 | 0·536 | −2·17* |
| Clinical follow-up; n = 7,469 | k-UPBioAge | 7·93 | 10·21 | 0·808 | 0·554 |
| | UPP-age | 9·87 | 12·67 | 0·655 | 0·313 |
| FLEMENGHO; n = 778 | k-UPBioAge | 5·44 | 7·15 | 0·894 | 0·796 |
| | UPP-age | 7·44 | 9·63 | 0·822 | 0·629 |

| All-cause mortality (n = 7,469; 625 deaths) | Measure | HR per SD (95% CI) |
|---|---|---|
| Age and sex | k-UPBioAgeAcc | 1·62 (1·49–1·76) |
| Age and sex | UPP-age acceleration | 1·33 (1·24–1·43) |
| Fully adjusted | k-UPBioAgeAcc | 1·47 (1·34–1·62) |
| Fully adjusted | UPP-age acceleration | 1·24 (1·15–1·33) |

One row is one clock evaluated in one cohort. k-UPBioAge is the kidney-conditioned prediction produced by Filtrate-Aware Calibration; UPP-age is the published Martens et al.[10] urinary peptidomic age predictor (54 peptides from 17 proteins), applied without re-fitting and without an age-bias correction. Both were applied to identical samples, so the comparison is matched. MAE, mean absolute error (years); RMSE, root mean squared error (years); r, Pearson correlation between predicted and chronological age; $R^2$, coefficient of determination, computed against each cohort's own age variance. Lower MAE and RMSE are better; higher r and $R^2$ are better. * $R^2$ is negative in the Glasgow cohort because its restricted age range makes squared prediction error exceed the error of predicting that cohort's mean age; the value is reported rather than suppressed, and reflects absolute calibration rather than ranking. Because FLEMENGHO forms part of UPP-age's original training set but was held out from the present clock, that row is conservative in UPP-age's favour. The lower block reports the association of each clock's age acceleration with all-cause mortality in the clinical follow-up cohort, per standard deviation of that measure (k-UPBioAgeAcc SD 9·3 years; UPP-age acceleration SD 11·5 years), under identical adjustment; excluding the 998 participants of the EPOGH and Heart failure studies, which include UPP-age's FLEMENGHO training participants (n = 6,471; 544 deaths), the fully adjusted HRs were 1·36 (95% CI 1·23–1·50) for k-UPBioAgeAcc and 1·21 (1·12–1·31) for UPP-age acceleration; the fully adjusted model includes age, sex, diabetes, cardiovascular disease, hypertension, body-mass index, eGFR, and mean arterial pressure. eGFR, estimated glomerular filtration rate; FLEMENGHO, Flemish Study on Environment, Genes and Health Outcomes; MAE, mean absolute error; SD, standard deviation; k-UPBioAge, kidney-conditioned urinary peptidomic biological age; k-UPBioAgeAcc, kidney-conditioned urinary peptidomic age acceleration; UPP-age, urinary peptidomic profiling age.

**Supplementary Table S14 | Mortality association of k-UPBioAgeAcc according to whether a Filtrate-Aware correction was applied, and according to whether albuminuria was measured or estimated.**

| Subset | n/N (%) | Age and sex HR (95% CI) | Fully adjusted HR (95% CI) |
|---|---|---|---|
| **Filtrate-Aware correction (fully adjusted includes CVD)** | | | |
| **Full cohort** | 625/7,469 (8·4%) | 1·62 (1·49–1·76) | 1·47 (1·34–1·62) |
| **No correction applied** | 118/3,649 (3·2%) | 1·23 (0·95–1·59) | 1·33 (1·02–1·74) |
| **Correction applied** | 507/3,820 (13·3%) | 1·45 (1·31–1·60) | 1·38 (1·24–1·53) |
| **P-interaction** | | 0·21 | 0·20 |
| **Albuminuria (fully adjusted excludes CVD; see note)** | | | |
| **Full cohort** | 625/7,469 (8·4%) | 1·62 (1·49–1·76) | 1·57 (1·43–1·72) |
| **Measured** | 200/2,482 (8·1%) | 1·71 (1·45–2·00) | 1·79 (1·52–2·11) |
| **Estimated** | 425/4,987 (8·5%) | 1·62 (1·47–1·80) | 1·35 (1·21–1·52) |
| **P-interaction** | | 0·79 | 0·39 |

HR (95% CI) for all-cause mortality per SD increment in k-UPBioAgeAcc, on the SD of the full clinical follow-up cohort so that every row is directly comparable; n = deaths and N = participants at risk. Age and sex: adjusted for age and sex. Fully adjusted: adjusted for age, sex, diabetes, cardiovascular disease, hypertension, body-mass index, measured eGFR, and mean arterial pressure; in the albuminuria block it omits cardiovascular disease, which is too closely tied to death in the measured-albuminuria subset for the model to converge. The first block separates the participants whose prediction the Filtrate-Aware Calibration left unchanged from those it corrected; the second separates those with a measured urinary albumin-to-creatinine ratio from those without. The two albuminuria groups are not exchangeable: the measured group is younger (mean age 48·7 versus 61·2 years) and has better filtration (mean eGFR 97·7 versus 69·5 mL/min/1·73 m²). P-interaction is a likelihood-ratio test of the product of k-UPBioAgeAcc and the grouping variable, fitted in the whole cohort.

**Supplementary Data**

Supplementary Data 1 | Peptide-level model detail. Share of total model weight and parent protein for each of the 2,631 peptides retained in the UPBioAge clock, supplied as a separate spreadsheet (SupplementaryData1_UPBioAge_peptides.xlsx). The signed Stage-2 coefficients and the complete fitted model are in the Zenodo deposit (doi:10.5281/zenodo.22914811). eGFR, estimated glomerular filtration rate; HR, hazard ratio; SD, standard deviation; UPBioAge, urinary peptidomic biological age.

**References**


1 Keller F, Beige J, Siwy J, *et al.* Urinary peptides provide information about the risk of mortality across a spectrum of diseases and scenarios. *J Transl Med* 2023; **21**: 663.

2 Frantzi M, Keller F, Latosinska A, *et al.* Urinary Collagen Peptides Predict Mortality. *Proteomics* 2026; **26**: 98–110.

3 Mavrogeorgis E, Mischak H, Latosinska A, Siwy J, Jankowski V, Jankowski J. Reproducibility Evaluation of Urinary Peptide Detection Using CE-MS. *Molecules* 2021; **26**: 7260.

4 Latosinska A, Siwy J, Mischak H, Frantzi M. Peptidomics and proteomics based on CE-MS as a robust tool in clinical application: The past, the present, and the future. *Electrophoresis* 2019; **40**: 2294–308.

5 Klein J, Papadopoulos T, Mischak H, Mullen W. Comparison of CE-MS/MS and LC-MS/MS sequencing demonstrates significant complementarity in natural peptide identification in human urine. *Electrophoresis* 2014; **35**: 1060–4.

6 Shen X, Wang C, Zhou X, *et al.* Nonlinear dynamics of multi-omics profiles during human aging. *Nat Aging* 2024; **4**: 1619–34.

7 Lu AT, Quach A, Wilson JG, *et al.* DNA methylation GrimAge strongly predicts lifespan and healthspan. *Aging* 2019; **11**: 303–27.

8 Higgins-Chen AT, Thrush KL, Wang Y, *et al.* A computational solution for bolstering reliability of epigenetic clocks: implications for clinical trials and longitudinal tracking. *Nat Aging* 2022; **2**: 644–61.

9 Le TT, Kuplicki RT, McKinney BA, *et al.* A Nonlinear Simulation Framework Supports Adjusting for Age When Analyzing BrainAGE. *Front Aging Neurosci* 2018; **10**: 317.

10 Martens DS, Thijs L, Latosinska A, *et al.* Urinary peptidomic profiles to address age-related disabilities: a prospective population study. *Lancet Healthy Longev* 2021; **2**: e690–703.

11 Wen J. Refining the generation, interpretation and application of multi-organ, multi-omics biological aging clocks. *Nat Aging* 2025; **5**: 1897–913.

12 Pencina MJ, D’Agostino RB, D’Agostino RB, Vasan RS. Evaluating the added predictive ability of a new marker: From area under the ROC curve to reclassification and beyond. *Stat Med* 2008; **27**: 157–72.

13 Pencina MJ, D’Agostino RB, Steyerberg EW. Extensions of net reclassification improvement calculations to measure usefulness of new biomarkers. *Stat Med* 2011; **30**: 11–21.

14 Jaimes Campos MA, Andújar I, Keller F, *et al.* Prognosis and Personalized In Silico Prediction of Treatment Efficacy in Cardiovascular and Chronic Kidney Disease: A Proof-of-Concept Study. *Pharmaceuticals* 2023; **16**: 1298.

15 Ferrucci L, Fabbri E. Inflammageing: chronic inflammation in ageing, cardiovascular disease, and frailty. *Nat Rev Cardiol* 2018; **15**: 505–22.

16 Fibrinogen Studies Collaboration. Plasma Fibrinogen Level and the Risk of Major Cardiovascular Diseases and Nonvascular Mortality. *JAMA* 2005; **294**: 1799–809.

17 Steubl D, Buzkova P, Garimella PS, *et al.* Association of serum uromodulin with mortality and cardiovascular disease in the elderly—the Cardiovascular Health Study. *Nephrol Dial Transplant* 2020; **35**: 1399–405.

18 Pivin E, Ponte B, de Seigneux S, *et al.* Uromodulin and Nephron Mass. *Clin J Am Soc Nephrol* 2018; **13**: 1556–7.

19 Baird DP, Reck M, Campbell R, *et al.* Urinary Clusterin is a Biomarker of Renal Epithelial Senescence and Predicts Human Kidney Disease Progression. *Kidney Int Rep* 2025; **10**: 2344–56.

20 Horvath S. DNA methylation age of human tissues and cell types. *Genome Biol* 2013; **14**: R115.

21 Argentieri MA, Xiao S, Bennett D, *et al.* Proteomic aging clock predicts mortality and risk of common age-related diseases in diverse populations. *Nat Med* 2024; **30**: 2450–60.

22 Chen BH, Marioni RE, Colicino E, *et al.* DNA methylation-based measures of biological age: meta-analysis predicting time to death. *Aging* 2016; **8**: 1844–65.

23 Xiao H, Lau C-HE, Dehghan A, Robinson O. Proteomic aging clocks in epidemiological studies: advances, applications and prospects. *Nat Aging* 2026; **6**: 970–86.

24 Havelka M, Satomura A, Yamaguchi H, *et al.* A urinary microRNA aging clock accurately predicts biological age. *Npj Aging* 2025; **12**: 14.

25 Ammous F, Zhao W, Ratliff SM, *et al.* Epigenetic age acceleration is associated with cardiometabolic risk factors and clinical cardiovascular disease risk scores in African Americans. *Clin Epigenetics* 2021; **13**: 55.

26 Zhang Z, Reynolds SR, Stolrow HG, Chen J, Christensen BC, Salas LA. Deciphering the role of immune cell composition in epigenetic age acceleration: Insights from cell-type deconvolution applied to human blood epigenetic clocks. *Aging Cell* 2024; **23**: e14071.

27 de Lange A-MG, Cole JH. Commentary: Correction procedures in brain-age prediction. *NeuroImage Clin* 2020; **26**: 102229.

28 Belczacka I, Latosinska A, Siwy J, *et al.* Urinary CE-MS peptide marker pattern for detection of solid tumors. *Sci Rep* 2018; **8**: 5227.

29 Van Calster B, McLernon DJ, van Smeden M, Wynants L, Steyerberg EW. Calibration: the Achilles heel of predictive analytics. *BMC Med* 2019; **17**: 230.

30 Latosinska A, Mina IK, Nguyen TMN, *et al.* In silico prediction of optimal multifactorial intervention in chronic kidney disease. *J Transl Med* 2025; **23**: 943.

31 Tofte N, Lindhardt M, Adamova K, *et al.* Early detection of diabetic kidney disease by urinary proteomics and subsequent intervention with spironolactone to delay progression (PRIORITY): a prospective observational study and embedded randomised placebo-controlled trial. *Lancet Diabetes Endocrinol* 2020; **8**: 301–12.

32 Lindhardt M, Persson F, Zürbig P, *et al.* Urinary proteomics predict onset of microalbuminuria in normoalbuminuric type 2 diabetic patients, a sub-study of the DIRECT-Protect 2 study. *Nephrol Dial Transplant* 2017; **32**: 1866–73.

33 Packham DK, Wolfe R, Reutens AT, *et al.* Sulodexide Fails to Demonstrate Renoprotection in Overt Type 2 Diabetic Nephropathy. *J Am Soc Nephrol* 2012; **23**: 123–30.

34 Verbeke F, Siwy J, Van Biesen W, *et al.* The urinary proteomics classifier chronic kidney disease 273 predicts cardiovascular outcome in patients with chronic kidney disease. *Nephrol Dial Transplant* 2021; **36**: 811–8.

35 Delles C, Schiffer E, von zur Muhlen C, *et al.* Urinary proteomic diagnosis of coronary artery disease: identification and clinical validation in 623 individuals. *J Hypertens* 2010; **28**: 2316–22.

36 Huang Q-F, Trenson S, Zhang Z-Y, *et al.* Urinary Proteomics in Predicting Heart Transplantation Outcomes (uPROPHET)—Rationale and database description. *PLoS One* 2017; **12**: e0184443.

37 He T, Melgarejo JD, Clark AL, *et al.* Serum and urinary biomarkers of collagen type-I turnover predict prognosis in patients with heart failure. *Clin Transl Med* 2021; **11**: e267.

38 Rudnicki M, Siwy J, Wendt R, *et al.* Urine proteomics for prediction of disease progression in patients with IgA nephropathy. *Nephrol Dial Transplant* 2021; **37**: 42–52.

39 Htun NM, Magliano DJ, Zhang Z-Y, *et al.* Prediction of acute coronary syndromes by urinary proteome analysis. *PLoS One* 2017; **12**: e0172036.

40 Frantzi M, van Kessel KE, Zwarthoff EC, *et al.* Development and Validation of Urine-based Peptide Biomarker Panels for Detecting Bladder Cancer in a Multi-center Study. *Clin Cancer Res* 2016; **22**: 4077–86.

41 Rossing K, Bosselmann HS, Gustafsson F, *et al.* Urinary Proteomics Pilot Study for Biomarker Discovery and Diagnosis in Heart Failure with Reduced Ejection Fraction. *PLoS One* 2016; **11**: e0157167.

42 Rauen T, Eitner F, Fitzner C, *et al.* Intensive Supportive Care plus Immunosuppression in IgA Nephropathy. *N Engl J Med* 2015; **373**: 2225–36.

43 Kuznetsova T, Mischak H, Mullen W, Staessen JA. Urinary proteome analysis in hypertensive patients with left ventricular diastolic dysfunction. *Eur Heart J* 2012; **33**: 2342–50.

44 Alkhalaf A, Zürbig P, Bakker SJL, *et al.* Multicentric Validation of Proteomic Biomarkers in Urine Specific for Diabetic Nephropathy. *PLoS One* 2010; **5**: e13421.
